# No convergence in three decades: national trajectories of episode-fatality ratios for childhood lower respiratory infections in 204 countries, 1990-2023

**DOI:** 10.64898/2026.09.01.26361942

**Authors:** Deze Li, Hao Chen, Xiaotong Wang, Chen Shen

**Affiliations:** Beijing Pediatric Research Institute, Beijing Children’s Hospital, Capital Medical University, Beijing, China

**Keywords:** lower respiratory infection, episode-fatality ratio, convergence, survival frontier, joinpoint regression, Global Burden of Disease

## Abstract

**Background:** Lower respiratory infections (LRI) remain the leading infectious cause of death in children, and survival once ill is a direct tracer of health-system quality. Whether countries are converging toward the best survival performance achieved within their own region has never been tested at national level. We measured each country’s distance to an empirical episode-fatality-ratio (EFR) frontier in 204 countries from 1990 to 2023.

**Methods:** For each country and year we computed EFR = LRI deaths/incident episodes using Global Burden of Disease (GBD) 2023 estimates for ages 0–19 years. Deaths span the full 1990–2023 series; episodes are observed for 1990, 2019 and 2023, with intermediate years linearly interpolated. The frontier was the 10th-percentile country EFR within each GBD super-region and year (sensitivity: 5th and 25th percentiles); the gap = EFR_country/EFR_frontier. We classified 33-year gap trajectories into catch-up phenotypes, ranked COVID-window (2019–2023) movers, cross-tabulated gap against avoidable deaths to build a priority list, and benchmarked upper respiratory infections (URI) at three time points as a near-zero-fatality contrast.

**Findings:** The median country’s gap was 1.86 in 1990, 1.80 in 2019 and 1.86 in 2023; the share of countries more than twice their regional frontier was 44.6% in 1990 and 46.6% in 2023. Of 137 eligible countries, 67 narrowed and 69 widened their gap, with one unchanged. Nineteen countries achieved sustained catch-up, concentrated in North Africa and the Middle East (7) and Latin America (5), with China closing from 2.43 to 0.50, below its regional frontier; 28 countries regressed, led by Central Asia (Uzbekistan ×3.5) and including the United States (×2.0). Over the COVID-19 window the median gap peaked at 2.00 in 2021 (+10.8% versus 2019, from unrounded medians) before returning to 1.86. Combining gap with avoidable deaths identifies two distinct policy problems: high-burden, moderate-gap giants (Nigeria 67,490 avoidable deaths, gap 2.4; India 54,109, gap 1.6) and extreme-gap outliers (Uzbekistan, gap 28.6). The Sub-Saharan Africa frontier fell further behind the High-income frontier (ratio 4.2 in 1990, 9.5 in 2023); the median Sub-Saharan African country sits 11.0 times the global 10th-percentile frontier but only 1.78 times its own regional frontier, so within-region benchmarking understates the region’s true distance. URI gaps likewise did not converge (median 4.15 to 4.60). Joinpoint regression formalized the pattern: the median gap showed no net narrowing over 33 years (average annual percent change +0.03% per year, 95% CI −0.05 to 0.12).

**Interpretation:** Convergence toward the survival frontier is not the default national trajectory: over three decades the typical country made no net progress toward the best decile of its own region, and pandemic-era divergence was only partly reversed. National gap trajectories separate system-wide quality shortfalls from extreme outliers warranting audit, and expose a measurement trap in which regions whose frontiers stagnate appear closer to best practice than they are.

## Introduction

Lower respiratory infections (LRI) remain the leading infectious cause of death among children and adolescents. Global Burden of Disease (GBD) 2023 estimates attribute 711,228 deaths among 0–19-year-olds to LRI in 2023, down from 2,035,714 in 1990, a 65% decline driven jointly by falling incidence and improving survival once infected.^1–3^ The two components have not contributed equally: incident episodes roughly halved over the period, while the probability of dying once ill fell by about 28%, so most of the mortality decline reflects fewer children falling ill rather than substantially better survival among the ill.^1,2^ The clinical pathway from pneumonia episode to death is short, well understood and responsive to inexpensive interventions (antibiotics, oxygen and supportive care) that have been standard of care for decades.^4^ Where health systems function, children should not die of pneumonia; the condition therefore serves as a tracer for the proposition that treatable deaths signal remediable system performance.

That proposition has a long intellectual lineage. Rutstein and colleagues proposed in 1976 that deaths from listed conditions be counted as sentinel events of health-care quality, each one a possible failure of prevention or treatment warranting investigation.^5^ Nolte and McKee developed the idea into the amenable-mortality framework that now underpins cross-national health-system comparison,^6^ and the Lancet Global Health Commission on High Quality Health Systems estimated that 8.6 million deaths annually in low– and middle-income countries are amenable to health care, with poor-quality care implicated in more deaths than non-use of services.^7^ The GBD Healthcare Access and Quality (HAQ) Index operationalized the concept by scoring each country against the lowest observed age-specific death rates for 32 causes, an empirical frontier of attained performance.^8^ These frameworks share a structural limitation for respiratory infections: they benchmark population-level mortality, conflating how many children fall ill with how many die once ill. A country with many episodes records more amenable deaths than one with few, even if both treat each episode equally well.

The episode-fatality ratio (EFR), deaths divided by incident episodes, separates these two quantities. It isolates the component of mortality most plausibly responsive to case management, and it can be computed annually for every country from standard GBD outputs without external development covariates. An EFR frontier, defined as the 10th-percentile country EFR within each GBD super-region and year, sizes that opportunity directly: benchmarked against within-region best practice, 333,803 childhood LRI deaths in 2023 (46.9% of the total) were avoidable, and the avoidable share has not narrowed since 1990. A frontier establishes the size of the opportunity at regional level; it does not ask whether individual countries are closing on the frontier over time. Regional aggregates can conceal national heterogeneity in both directions: a stagnating regional average is compatible with some countries catching up and others falling back, and the policy response differs fundamentally between a region of uniform laggards and one of divergent movers. Resolving which pattern holds requires country-year resolution.

This is the question the Sustainable Development Goals (SDG) era takes for granted. The default narrative of global child health is one of convergence: interventions diffuse, coverage rises, and lagging countries catch up with the best performers.^9^ The narrative has empirical support in some domains. Gaps in amenable mortality between European countries narrowed over recent decades as effective care diffused eastward,^6^ and global declines in under-five mortality have been fastest in many high-burden settings.^9^ But mortality declines are consistent with convergence without demonstrating it: deaths can fall because fewer children fall ill while each country’s relative position against its regional frontier stays fixed. Frontier-style benchmarking has been applied to national mortality levels,^8,10^ and formal convergence tests on mortality itself have already returned negative verdicts: child mortality indicators showed β– and σ-divergence across 187 countries over 1990–2010,^11^ and a global analysis of nine mortality indicators from 1990 to 2030 found convergence clubs rather than universal convergence.^12^ Our increment is specific: to our knowledge no study has tested, country by country and year by year, whether national case-survival performance, net of incidence, is converging toward an empirically attained frontier for a major treatable cause of child death. The distinction is not academic. If convergence is underway, the residual burden is a matter of time and sustained financing; if it is not, the machinery assumed to diffuse best practice (guidelines, commodities, training and financing flows) is failing in ways that aggregate mortality trends cannot reveal.

This study provides that test for childhood LRI in 204 countries from 1990 to 2023. We ask three questions. First, has the distance between the typical country and its regional survival frontier narrowed over three decades, globally and within each super-region? Second, which countries sustained catch-up, which stalled and which regressed, where are they concentrated, and how did national gaps move over the 2019–2023 pandemic window? Third, where do large gaps coincide with large avoidable burdens, and does benchmarking within regions systematically understate the distance for regions whose frontiers have themselves stagnated? The answers determine whether the residual burden of a treatable killer is a problem of catching up, of advancing the frontier, or of both, and for whom.

## Methods

### Study design and data sources

This is a country-level trend analysis of modelled estimates from GBD 2023,^1,2^ which synthesizes vital registration, surveillance, verbal autopsy and survey data through standardized cause-of-death and non-fatal modelling to produce internally consistent estimates for all locations and years. We extracted LRI deaths and incident episodes for ages 0–19 years (GBD age groups under 5, 5–9, 10–14 and 15–19 years, summed) for 204 countries and territories. LRI deaths span the full 1990–2023 annual series for all 204 countries; country sums reproduce published GBD totals (2,035,714 deaths in 1990; 711,228 in 2023).^1^ Country-level LRI episodes are observed for three anchor years, 1990, 2019 and 2023, spanning the MDG-era baseline, the pre-pandemic reference year and the most recent GBD cycle; episodes for intermediate years were linearly interpolated between the observed anchors. This interpolation is a declared approximation: episodes move smoothly relative to deaths, so annual gap dynamics are dominated by the observed death series, and all anchor-year EFRs, gaps and phenotype classifications rely on observed inputs only. The smoothing assumption is most exposed over the 2020–2022 pandemic window, when respiratory incidence departed sharply from smooth trends; annual gap values for those years, including the 2021 peak, therefore rest on interpolated denominators and should be read as smoothed approximations, whereas comparisons between observed anchors (2019 versus 2023) do not. As a secondary cross-check we assembled an upper respiratory infection (URI) panel of deaths and episodes for 1990, 2019 and 2023 with no interpolation. Countries were assigned to the seven GBD 2023 super-regions (5–46 countries each). All inputs are publicly released modelled aggregates; no individual-level data were used and ethics review was not required.

### Episode-fatality ratio, frontier and gap

For country i in year y, EFR = LRI deaths/incident episodes. The EFR is not a clinical case-fatality ratio: GBD episode denominators include mild community and outpatient illness (81.0 million LRI episodes globally in 2023), so EFRs lie far below facility-based fatality rates and summarize, for an entire national system, the probability that an episode ends in death.^2,4^ The mortality-to-incidence ratio has a long history as a system-level survival proxy in cancer epidemiology, where its validity has been debated because deaths and cases arise from different cohorts;^13^ for acute respiratory infections, whose course from onset to death is measured in days to weeks, that objection carries little force.

The frontier EFR is the 10th percentile of country EFRs within each super-region × year stratum. Benchmarking within super-regions requires no external income or development covariate, avoids adjusting a performance measure by a covariate that is itself partly a function of health-system output, and respects broad regional epidemiological contexts such as pathogen mix and seasonality. The 10th percentile, rather than the minimum, prevents any single country estimate, potentially distorted by small numbers or model artefacts at the favourable extreme, from defining best practice for an entire region. The rationale holds only partially in South Asia, whose five countries place the interpolated 10th percentile at or near the best-performing country, so the frontier there approximates a best-country rather than a best-decile definition. The frontier is recomputed annually, so benchmarks evolve with observed performance.

The gap is gap_i = EFR_i/EFR_frontier,sr(i). A gap of 1 places a country at its regional frontier; a gap of 3 means its children die once ill at three times the rate of the best-performing decile of its own region. Avoidable deaths are defined as avoidable_i = max(0, deaths_i − episodes_i × EFR_frontier,sr(i)); applied to the 2023 national panel this reproduces the global LRI avoidable total of 333,803 exactly. Because the frontier is relative, gaps understate absolute distance from global best practice wherever an entire region lags, a point we quantify directly for Sub-Saharan Africa.

### Catch-up phenotypes

Each eligible country’s trajectory was classified from gap anchors at 1990, 2010, 2019 and 2023. Countries with fewer than 25 LRI deaths in 2023 were set aside as low-count (ratios unstable; excluded from rankings but retained in the panel). The threshold is applied to 2023, the ranking year; of the 67 countries below it, 37 were also below it in 1990, while 30 countries—the largest group high-income (12 of 30)—whose death counts fell under 25 as mortality declined, are excluded from classification despite having informative long-run trajectories. Among the remainder: near-frontier countries held a gap of 1.5 or less at all four anchors; sustained catch-up requires gap_2023/gap_1990 ≤ 0.7 (at least 30% relative closure) with no net widening by 2019; regressing requires gap_2023/gap_1990 ≥ 1.3; all remaining eligible countries are classified as stalled. COVID-window movers were ranked by gap_2023/gap_2019 among eligible countries. Thresholds (±30%, 1.5, 25 deaths) are conventional; the full country listing is published so alternatives can be re-derived. Because a fixed ±30% relative-change threshold is mechanically easier to satisfy from an extreme 1990 gap, the catch-up and regressing classes will partly reflect regression to the mean; the formal β-convergence statistic reported in the Results quantifies this tendency directly.

### Priority grid and regional benchmark diagnostics

To locate where catch-up effort buys the most lives, we cross-tabulated each country’s 2023 gap against its absolute avoidable deaths and ranked countries by a gap-weighted burden, avoidable deaths × ln(gap), which rises with both the size of the avoidable toll and the proportional distance from the frontier. To diagnose benchmark degradation, we computed the ratio of the Sub-Saharan Africa frontier to the High-income frontier in 1990 and 2023, counted Sub-Saharan African countries at or near their own frontier (gap ≤ 1.5) at each anchor, and compared the median Sub-Saharan African country’s gap to its own regional frontier with its gap to the global 10th-percentile frontier (1.12 per 1,000 episodes in 2023), the latter approximating distance from achievable global best practice.

### Sensitivity analyses and uncertainty

The primary frontier percentile was varied (5th and 25th versus 10th) and all headline patterns recomputed. The URI panel bounds the method at the near-zero-fatality extreme without interpolation. All inputs are GBD 2023 modelled estimates rather than vital-registration counts, and national estimates carry the uncertainty of the underlying GBD models; primary estimates here are deterministic, computed from GBD central values consistent with published sums. Two external checks use data outside the GBD inputs: we correlated the 2023 gap with national mean elevation, computed from the NOAA ETOPO1 global relief model,^14^ and with the WHO/World Bank UHC service coverage index^15^ (Spearman rank correlations). We do not adjust for national ethnic composition (no comparable country-level data; no direct causal pathway to episode survival) or for coastal and ocean-climate variables (partially absorbed by super-region stratification). The main threats to inference are the declared episode interpolation, ratio instability at low death counts (addressed by the 25-death exclusion) and possible data-model artefacts behind extreme gaps in countries with weak vital registration, which we flag for audit rather than interpret literally.

### Joinpoint analysis of trajectories

To locate turning points in the aggregate trajectories, we fitted joinpoint regression—continuous (hinge) segments on the natural-log scale—to three annual series for 1990–2023: the global EFR (total LRI deaths divided by total episodes), the global median country gap, and the ratio of the Sub-Saharan Africa frontier EFR to the High-income frontier EFR. The number of joinpoints (zero to three) was selected by grid search minimizing the Bayesian information criterion, counting 2k + 2 parameters for a model with k joinpoints (k estimated joinpoint locations plus k + 2 regression coefficients), with at least three annual observations per segment and adjacent joinpoints constrained to lie at least three years apart; confidence intervals use n − (2k + 2) residual degrees of freedom. We report the annual percent change (APC) per segment with t-based 95% confidence intervals on the log scale and the span-weighted average annual percent change (AAPC) for 1990–2023 with delta-method intervals. Because episode denominators for intermediate years are interpolated, within-window movements of the EFR series reflect both the observed death series and the functional form of the interpolation; the denominator’s contribution is quantified segment by segment in the note to Table 6. Model selection remained sensitive to the interpolation of the episode denominators; we therefore report the AAPC as the primary summary, and the fits selected under the previous parameter counting are retained in the supplementary joinpoint CSV for comparison.

## Results

### The median country did not converge

Across 204 countries, the median gap to the within-super-region frontier was 1.86 in 1990, 1.80 in 2019 (−3.0%) and 1.86 in 2023: over 33 years the typical country ended exactly where it began relative to the best decile of its own region (Figure 1, Table 1). The share of countries more than twice their frontier was 44.6% in 1990, 44.1% in 2019 and 46.6% in 2023. Among the 137 countries eligible for classification, 67 narrowed and 69 widened their gap between 1990 and 2023, with one unchanged (the United Arab Emirates, at its regional frontier in both years): an almost even split; sustained catch-up was achieved by fewer than one in seven eligible countries (19 of 137).

**Figure 1.**
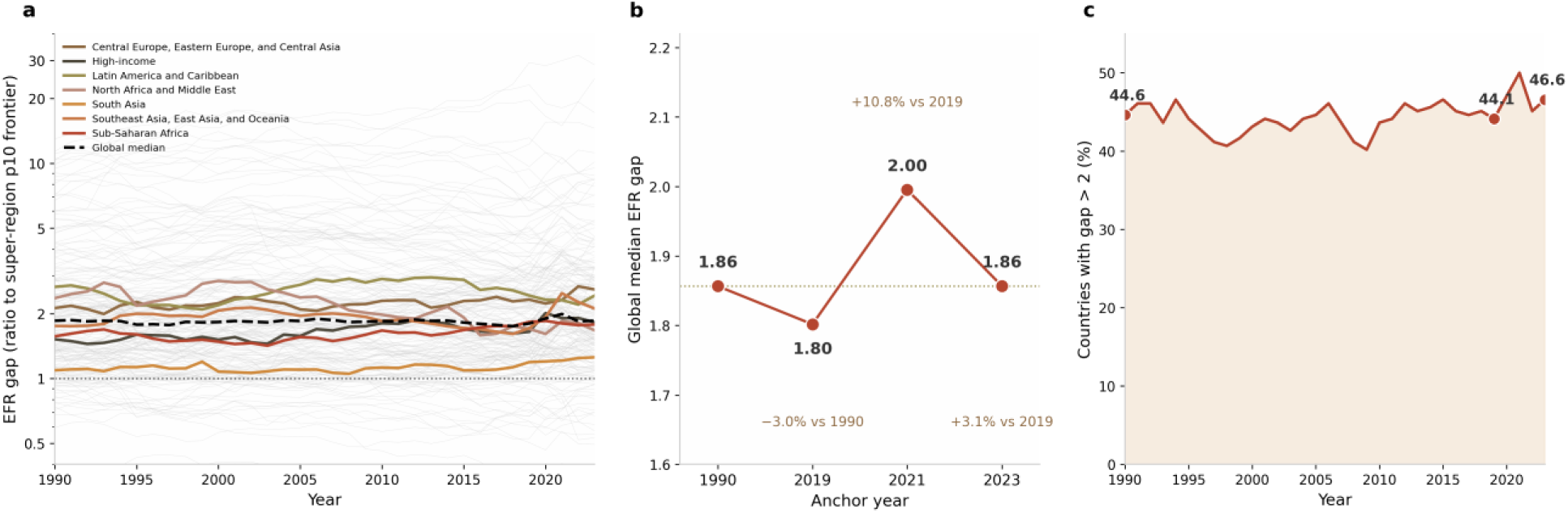
Distance to the LRI survival frontier, 204 countries, 1990–2023. (a) Country EFR gap trajectories (thin grey lines; country EFR / within-super-region 10th-percentile frontier EFR, log scale), super-region medians (thick coloured lines), and the global median (dashed line); the dotted line marks a gap of 1 (at the frontier). (b) Global median EFR gap at the anchor years: 1.86 (1990), 1.80 (2019), 2.00 (2021), 1.86 (2023); corresponding changes −3.0% (1990 to 2019), +10.8% (2019 to 2021), +3.1% (2019 to 2023). (c) Share of countries with gap > 2 by year: 44.6% (1990), 44.1% (2019), 46.6% (2023).

**Table 1.** Median EFR gap to the within-super-region LRI survival frontier, by super-region and year (ages 0–19 years)

| **Super-region** | **1990** | **2010** | **2019** | **2021** | **2023** |
|---|---|---|---|---|---|
| Central Europe, Eastern Europe, and Central Asia | 2.13 | 2.30 | 2.33 | 2.32 | 2.60 |
| High-income | 1.52 | 1.81 | 1.65 | 1.91 | 1.82 |
| Latin America and Caribbean | 2.67 | 2.90 | 2.43 | 2.32 | 2.43 |
| North Africa and Middle East | 2.36 | 1.98 | 1.70 | 1.85 | 1.68 |
| South Asia* | 1.09 | 1.12 | 1.19 | 1.21 | 1.25 |
| Southeast Asia, East Asia, and Oceania | 1.76 | 1.85 | 1.72 | 2.49 | 2.12 |
| Sub-Saharan Africa | 1.57 | 1.67 | 1.83 | 1.80 | 1.78 |
| Global median | 1.86 | 1.85 | 1.80 | 2.00 | 1.86 |
| Countries with gap >2 (%) | 44.6 | – | 44.1 | – | 46.6 |

EFR, episode-fatality ratio (deaths per incident episode). Gap = country EFR/frontier EFR, where the frontier is the 10th percentile of country EFRs within each super-region and year; a gap of 1 places a country at its regional frontier. \*South Asia contains five countries, so its 10th-percentile frontier approximates a best-country definition. The 2021 column relies in part on linearly interpolated episode denominators and is a smoothed approximation (see Methods).

The absence of convergence is visible throughout the series. The global median gap in 2010 was 1.85, essentially unchanged from 1990 (1.86), 2019 (1.80) and 2023 (1.86): three decades of expanding intervention coverage left the typical country’s relative position untouched. Absolute performance did improve: national EFRs fell in 187 of 204 countries (92%) between 1990 and 2023; it is each country’s position relative to the moving regional frontier that did not advance.

### Joinpoint turning points of the EFR and gap trajectories

Joinpoint regression of the global median-gap trajectory selected no joinpoints (k = 0; Table 6): a single log-linear trend fitted over 1990–2023 gives an average annual percent change of +0.03% per year (95% CI −0.05 to 0.12)—a formal, interval-quantified confirmation that the typical country’s distance to its regional frontier did not narrow. The three-joinpoint fit obtained under the previous BIC parameter counting (joinpoints 2014, 2017 and 2020) is retained in the supplementary CSV for comparison; its 33-year summary is likewise statistically indistinguishable from zero. Sensitivity to this constraint choice: under the looser observation-count-only rule (no span requirement), the grid search would select k = 3 (joinpoints 2014, 2018, 2021; ΔBIC = −13.0 in favour of k = 3), whose span-weighted AAPC is +0.005% per year (95% CI −0.093 to +0.104)—equally indistinguishable from zero, so the non-convergence conclusion is invariant to the constraint choice.

The apparent mid-period rise in the global EFR is an interpolation artefact rather than a reversal of survival improvement. The global EFR fell from 12.26 to 8.78 deaths per 1,000 episodes over 1990–2023 (AAPC −1.33% per year, −1.54 to −1.12), with two joinpoints selected (2010 and 2017): slow improvement from 1990 to 2010 (APC −1.02%, −1.31 to −0.73), a fitted rise between 2010 and 2017 (APC +1.80%, 0.91 to 2.70) and a steep decline thereafter (2017–2023 APC −5.88%, −7.19 to −4.55). The 2010–2017 rise should not be read as a reversal of survival improvement: observed deaths fell monotonically throughout 2010–2017 (−14.7% cumulatively; −12.3% over 2011–2017), 61.7% of the segment’s log-scale EFR movement comes from the interpolated episode denominator rather than from deaths (Table 6 note), and the level-linear interpolation mechanically accelerates on the log scale, from −3.3% per year in 2011 to −4.1% per year by 2017. Under log-linear interpolation of the same episode anchors the cumulative 2011–2017 rise shrinks from +9.7% to +4.4% and is no longer monotone (the series peaks in 2016). The Sub-Saharan Africa-to-High-income frontier ratio rose from 4.24 to 9.53 between 1990 and 2023 (fitted AAPC +2.80% per year, 2.56 to 3.03), with joinpoints in 2004, 2014 and 2017, confirming that the regional frontiers themselves diverged even where within-region gaps were stable.

Super-region medians diverged rather than converged (Table 1). North Africa and the Middle East was the only region to converge strongly (median gap 2.36 to 1.68). South Asia remained closest to its frontier (1.09 to 1.25), although its five-country frontier approximates a best-country definition (Table 1). Two regions drifted away from their frontiers over the full period: Central Europe, Eastern Europe and Central Asia (2.13 to 2.60) and Southeast Asia, East Asia and Oceania (1.76 to 2.12). Latin America and the Caribbean narrowed modestly (2.67 to 2.43), while Sub-Saharan Africa (1.57 to 1.78) and the High-income super-region (1.52 to 1.82) drifted wider. The flat-median pattern is insensitive to the frontier definition: under 5th– and 25th-percentile frontiers the global median gap rose from 2.00 to 2.42 and from 1.37 to 1.43 respectively (Table 5).

Two formal convergence statistics quantify the same pattern (Supplementary Table S6). For σ-convergence, the cross-country coefficient of variation of the gap rose from 0.89 in 1990 to 1.18 in 2023, and the standard deviation of the log gap from 0.67 to 0.72: dispersion widened rather than narrowed. For β-convergence, regressing the 1990–2023 change in log gap on the 1990 log gap across the 137 eligible countries yields a slope of −0.10 (SE 0.05; r = −0.19, p = 0.03): countries with wider initial gaps did tend to narrow proportionally more, a weak catch-up tendency consistent in part with regression to the mean, but one far too weak to arrest the widening of the cross-country distribution. Convergence, where it operated at all, did so inside a distribution that was itself spreading out.

### Catch-up phenotypes and their geography

Of 204 countries, 63 were classified as stalled, 28 as regressing, 27 as near-frontier and 19 as sustained catch-up, with 67 low-count countries excluded from classification (Figure 3, Table 2). Sustained catch-up is geographically concentrated. Seven of the 19 catch-up countries are in North Africa and the Middle East (Türkiye, Iran, Sudan, Oman, Algeria, Syria, Yemen), five in Latin America (Bolivia, Peru, Honduras, Nicaragua, Costa Rica) and four in Central and Eastern Europe and Central Asia (Albania, Belarus, Mongolia, Serbia) (Table 2). The largest national catch-up is China’s: its gap fell from 2.43 to 0.50, placing it below its own regional frontier in 2023. Belarus (1.80 to 0.71) and the Republic of Korea (2.32 to 1.04) likewise closed to or below their frontiers. Equatorial Guinea is the only sustained catch-up country in Sub-Saharan Africa.

**Table 2.**
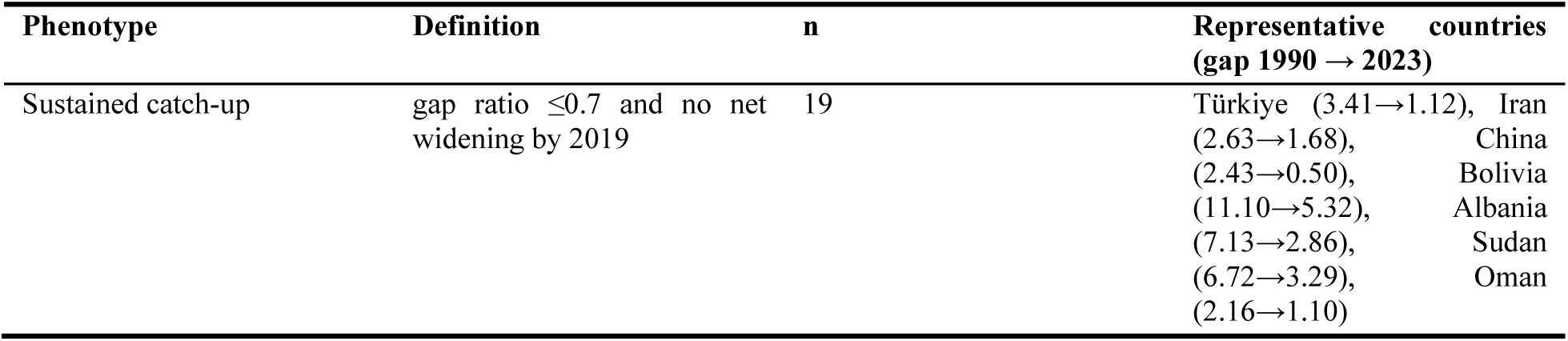
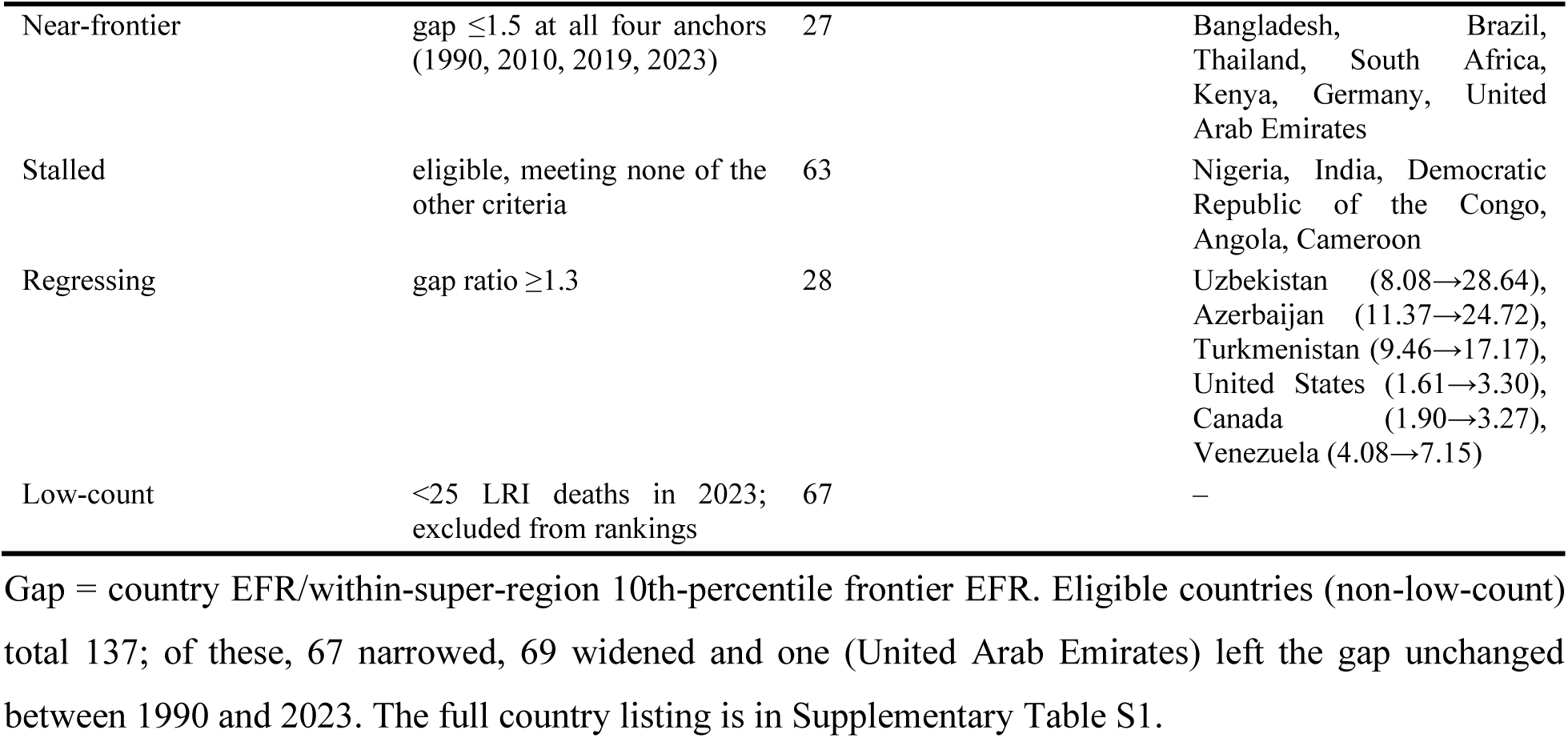
Catch-up phenotypes of 33-year gap trajectories, 1990–2023.

| Phenotype | Definition | n | Representative countries (gap 1990 → 2023) |
|---|---|---|---|
| Sustained catch-up | gap ratio ≤0.7 and no net widening by 2019 | 19 | Türkiye (3.41→1.12), Iran (2.63→1.68), China (2.43→0.50), Bolivia (11.10→5.32), Albania (7.13→2.86), Sudan (6.72→3.29), Oman (2.16→1.10) |
| Near-frontier | gap ≤1.5 at all four anchors (1990, 2010, 2019, 2023) | 27 | Bangladesh, Brazil, Thailand, South Africa, Kenya, Germany, United Arab Emirates |
| Stalled | eligible, meeting none of the other criteria | 63 | Nigeria, India, Democratic Republic of the Congo, Angola, Cameroon |
| Regressing | gap ratio ≥1.3 | 28 | Uzbekistan (8.08→28.64), Azerbaijan (11.37→24.72), Turkmenistan (9.46→17.17), United States (1.61→3.30), Canada (1.90→3.27), Venezuela (4.08→7.15) |
| Low-count | <25 LRI deaths in 2023; excluded from rankings | 67 | – |

Gap = country EFR/within-super-region 10th-percentile frontier EFR. Eligible countries (non-low-count) total 137; of these, 67 narrowed, 69 widened and one (United Arab Emirates) left the gap unchanged between 1990 and 2023. The full country listing is in Supplementary Table S1.

The 27 near-frontier countries include most of South Asia (Bangladesh, Bhutan, Nepal, Pakistan), Brazil and Cuba, Kenya, Uganda and South Africa, and Thailand and Malaysia; the group is geographically broad but demographically small relative to the stalled majority. The 63 stalled countries carry most of the global burden: Nigeria, India and the Democratic Republic of the Congo all sit in this class, with gaps that neither closed nor blew out over 33 years.

Regression is led by Central Asia: Uzbekistan’s gap multiplied 3.5-fold (8.08 to 28.64), Azerbaijan’s 2.2-fold (11.37 to 24.72) and Turkmenistan’s 1.8-fold (9.46 to 17.17), with Kazakhstan also regressing (5.98 to 8.44). High-income countries are not exempt: the United States doubled its gap (1.61 to 3.30), Canada widened 1.7-fold, and France, the United Kingdom, Argentina and Venezuela are also regressing. These high-income regressions are relative, not absolute: every named high-income regressor improved its own EFR over the period (the United States by about 36%, from 3.75 to 2.40 per 1,000 episodes), but the High-income frontier improved by about two-thirds, so distance to the frontier grew even as survival improved. Within Sub-Saharan Africa, 8 of 45 eligible countries are regressing against a single sustained catch-up. Eswatini illustrates the fragility of frontier membership: one of the five countries that defined the Sub-Saharan Africa frontier in 1990 (gap 0.86), it is classified as regressing in 2023 (gap 2.94).

### COVID-window divergence and its partial reversal

The pandemic window produced a sharp, largely transient divergence. The global median gap rose from 1.80 in 2019 to 1.90 in 2020 and 2.00 in 2021 (+10.8% versus 2019; percentages here and below are computed from unrounded medians), before falling back to 1.86 in 2023, a residual widening of +3.1% (Figure 1). The 2021 peak rests on interpolated episode denominators and is a smoothed estimate whose height and timing are sensitive to that assumption (see Methods); the observed-anchor comparison (2019 versus 2023, +3.1%) does not depend on it. National movements over the window were large in both directions (Figure 4, Table 3). The fastest catch-up was Botswana’s (gap 2.17 to 1.49), and the United Arab Emirates moved from 1.42 to 1.00, reaching its regional frontier; Japan was the only high-income country among the ten fastest catch-ups (Table 3). The largest regressions were Ecuador (+59%), Kazakhstan (+58%), Cambodia (+47%) and the United States (+44%), with France, Canada, Saudi Arabia and Peru each widening by a third or more. Afghanistan’s gap widened 36% to 6.3, against 10,268 LRI deaths in 2023: the window’s divergence hit hardest where the pre-pandemic gap was already wide. High-income countries appear on both sides of the ledger, reinforcing that pandemic-era disruption of case survival was not confined to low-income settings.

**Table 3.** COVID-window movers: fastest catch-up and largest regressions in EFR gap, 2019–2023 (eligible countries)

| **Catch-up (gap narrowed)** | **Super-region** | **Gap 2019** | **Gap 2023** | **Ratio** |
|---|---|---|---|---|
| Botswana | Sub-Saharan Africa | 2.17 | 1.49 | 0.69 |
| United Arab Emirates | North Africa and Middle East | 1.42 | 1.00 | 0.71 |
| Turkmenistan | Central Europe, Eastern Europe, and Central Asia | 23.13 | 17.17 | 0.74 |
| Albania | Central Europe, Eastern Europe, and Central Asia | 3.72 | 2.86 | 0.77 |
| Honduras | Latin America and Caribbean | 3.37 | 2.62 | 0.78 |
| Bolivia | Latin America and Caribbean | 6.66 | 5.32 | 0.80 |
| Gabon | Sub-Saharan Africa | 1.53 | 1.24 | 0.81 |
| El Salvador | Latin America and Caribbean | 4.28 | 3.56 | 0.83 |
| Japan | High-income | 1.38 | 1.15 | 0.83 |
| Slovakia | Central Europe, Eastern Europe, and Central Asia | 3.51 | 2.93 | 0.83 |
| Regressing (gap widened) |  |  |  |  |
| Ecuador | Latin America and Caribbean | 2.87 | 4.57 | 1.59 |
| Kazakhstan | Central Europe, Eastern Europe, and Central Asia | 5.33 | 8.44 | 1.58 |
| Cambodia | Southeast Asia, East Asia, and Oceania | 3.42 | 5.02 | 1.47 |
| United States | High-income | 2.30 | 3.30 | 1.44 |
| Georgia | Central Europe, Eastern Europe, and Central Asia | 3.11 | 4.36 | 1.40 |
| France | High-income | 1.10 | 1.54 | 1.40 |
| Canada | High-income | 2.35 | 3.27 | 1.39 |
| Afghanistan | North Africa and Middle East | 4.64 | 6.29 | 1.36 |
| Saudi Arabia | North Africa and Middle East | 1.89 | 2.54 | 1.34 |
| Peru | Latin America and Caribbean | 2.48 | 3.31 | 1.34 |

Ratio = gap 2023/gap 2019. Over the same window the global median gap rose from 1.80 (2019) to a peak of 2.00 (2021, +10.8%, computed from unrounded medians of 1.802 and 1.995) and returned to 1.86 (2023). The 2021 peak rests on interpolated episode denominators (see Methods).

### Where gaps meet burden: the priority list

The 2023 geography of the gap (Figure 2) shows extreme values clustered in Central Asia, with elevated gaps across much of Latin America, South and Southeast Asia and parts of Sub-Saharan Africa. Cross-tabulating the 2023 gap against absolute avoidable deaths separates two distinct policy problems (Table 4). The first is high-burden, moderate-gap giants: Nigeria (67,490 avoidable LRI deaths, gap 2.4), India (54,109, gap 1.6), Niger (18,738, gap 3.9), the Democratic Republic of the Congo (14,470, gap 1.9) and Chad (8,621, gap 2.6), where system-wide improvement in the quality of case management across very large episode volumes would avert the most deaths. The second is extreme-gap outliers: Uzbekistan (10,679 avoidable deaths against a gap of 28.6, the single most extreme value worldwide), Azerbaijan (24.7), Turkmenistan (17.2), Haiti (9.6) and Mexico (8.2), where fatality per episode so far exceeds regional best deciles that targeted review of case-management performance or data quality is warranted before programmatic inference. Afghanistan, Myanmar, Indonesia and Egypt combine substantial burdens with large gaps. The 15 countries in Table 4 together hold 229,230 avoidable deaths, 68.7% of the 2023 global total, so a bounded set of national programmes could in principle reach more than two-thirds of the avoidable burden.

**Figure 2.**
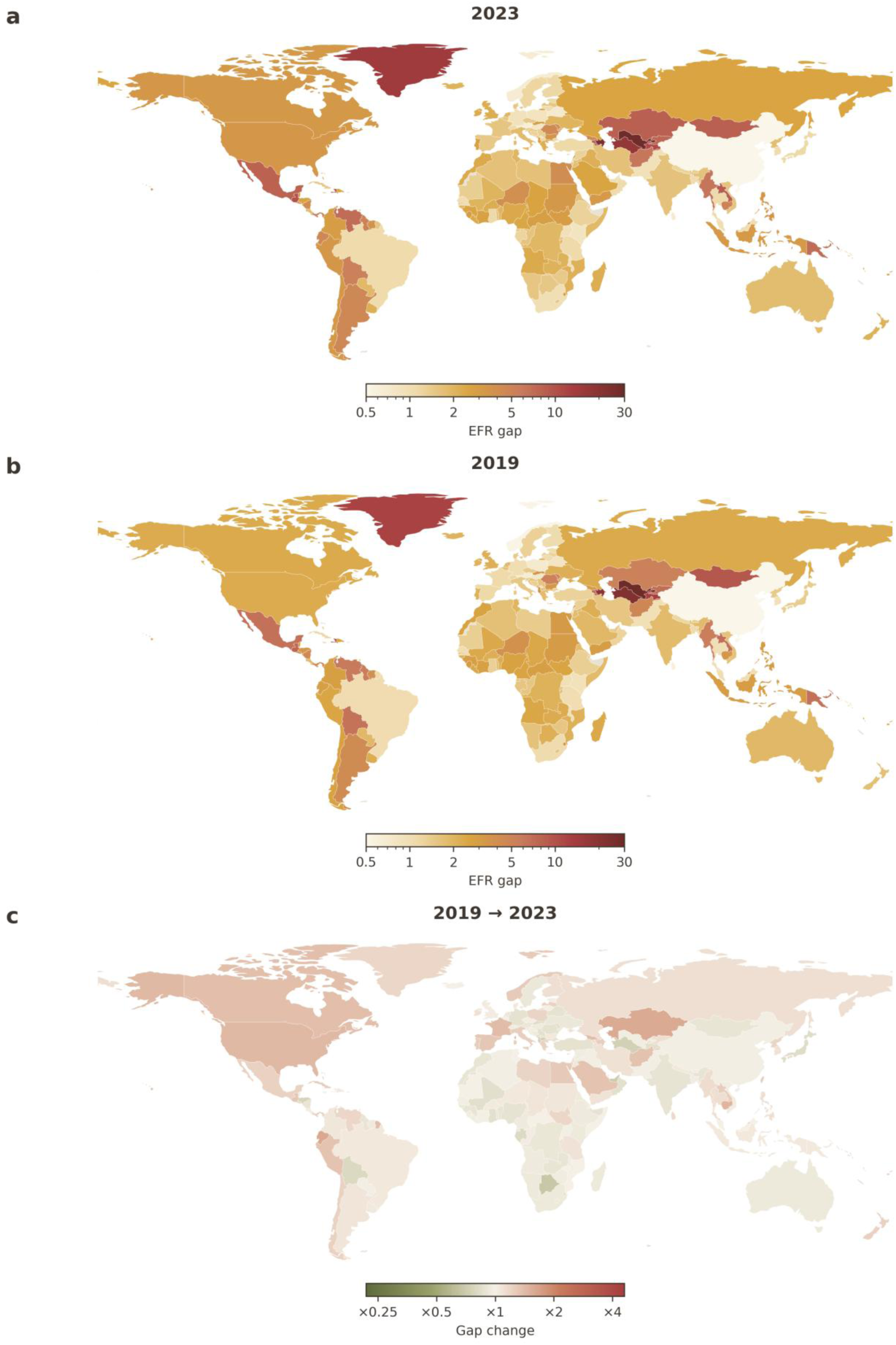
Distance to the LRI survival frontier by country (EFR ratio to within-super-region 10th-percentile frontier). (a) Gap in 2023 (log colour scale). (b) Gap in 2019 (same scale). (c) Fold change 2019 to 2023 (diverging scale centred on ×1; red = widening, olive = narrowing). Grey: no GBD 2023 estimate or not in the 1:110m base map.

**Table 4.** Priority countries, 2023: EFR gap × avoidable LRI deaths (top 15, ranked by avoidable deaths × ln gap)

| Country | Super-region | Gap 2023 | Avoidable deaths | Observed LRI deaths |
|---|---|---|---|---|
| Nigeria | Sub-Saharan Africa | 2.4 | 67,490 | 116,482 |
| Uzbekistan | Central Europe, Eastern Europe, and Central Asia | 28.6 | 10,679 | 11,065 |
| India | South Asia | 1.6 | 54,109 | 143,192 |
| Niger | Sub-Saharan Africa | 3.9 | 18,738 | 25,191 |
| Afghanistan | North Africa and Middle East | 6.3 | 8,634 | 10,268 |
| Myanmar | Southeast Asia, East Asia, and Oceania | 6.4 | 7,006 | 8,296 |
| Indonesia | Southeast Asia, East Asia, and Oceania | 3.2 | 9,757 | 14,197 |
| Egypt | North Africa and Middle East | 4.1 | 7,649 | 10,096 |
| Democratic Republic of the Congo | Sub-Saharan Africa | 1.9 | 14,470 | 30,177 |
| Chad | Sub-Saharan Africa | 2.6 | 8,621 | 13,920 |
| Philippines | Southeast Asia, East Asia, and Oceania | 3.4 | 6,223 | 8,767 |
| Haiti | Latin America and Caribbean | 9.6 | 3,379 | 3,774 |
| Côte d'Ivoire | Sub-Saharan Africa | 2.8 | 7,246 | 11,371 |
| Mexico | Latin America and Caribbean | 8.2 | 3,344 | 3,810 |
| Azerbaijan | Central Europe, Eastern Europe, and Central Asia | 24.7 | 1,884 | 1,964 |

Avoidable deaths = max(0, deaths − episodes × frontier EFR), deterministic estimates from GBD 2023 central values; the 2023 national panel sums to the global LRI avoidable total of 333,803. The ranking combines the size of the avoidable toll with the proportional distance from the frontier, separating high-burden, moderate-gap countries from extreme-gap outliers; ranks are computed on unrounded values. Displayed avoidable deaths sum to 229,229 owing to rounding; the unrounded total is 229,230 (68.7% of the global total).

**Table 5.** Sensitivity of the no-convergence finding to frontier definition, and URI cross-check (global median gap)

| Benchmark | 1990 | 2019 | 2023 |
|---|---|---|---|
| LRI, 5th-percentile frontier | 2.00 | 2.25 | 2.42 |
| LRI, 10th-percentile frontier (primary) | 1.86 | 1.80 | 1.86 |
| LRI, 25th-percentile frontier | 1.37 | 1.44 | 1.43 |
| URI, 10th-percentile frontier | 4.15 | 4.36 | 4.60 |

Under every frontier definition the median country's gap was as wide or wider in 2023 than in 1990. URI values derive from observed deaths and episodes at three time points, without interpolation. As a regional-benchmark diagnostic, the median Sub-Saharan African country's gap in 2023 was 1.78 to its own super-region frontier but 11.0 to the global 10th-percentile frontier (1.12 deaths per 1,000 episodes). In the URI panel, 93.5% of avoidable deaths in 2023 occurred in Sub-Saharan Africa.

**Table 6.** Joinpoint regression of the global EFR, median-gap and frontier-ratio trajectories, 1990– 2023.

| **Series (BIC-selected joinpoints)** | **Segment** | **APC, %/yr (95% CI)** | **AAPC 1990–2023, %/yr (95% CI)** |
|---|---|---|---|
| Global EFR, deaths per 1,000 episodes (2010, 2017) | 1990–2010 | −1.02 (−1.31, −0.73) | −1.33 (−1.54, −1.12) |
|  | 2010–2017 | +1.80 (0.91, 2.70) |  |
|  | 2017–2023 | −5.88 (−7.19, −4.55) |  |
| Global median gap to the within-super-region frontier (no joinpoints selected) | 1990–2023 | +0.03 (−0.05, 0.12) | +0.03 (−0.05, 0.12) |
| Sub-Saharan Africa / High-income frontier EFR ratio (2004, 2014, 2017) | 1990–2004 | +5.09 (4.59, 5.56) | +2.80 (2.56, 3.03) |
|  | 2004–2014 | +0.76 (0.09, 1.43) |  |
|  | 2014–2017 | −7.23 (−9.59, −4.81) |  |
|  | 2017–2023 | +6.28 (4.58, 8.01) |  |

Continuous hinge segments fitted to the natural log of each series by least squares; number of joinpoints (zero to three) selected by BIC grid search counting 2k + 2 parameters for k joinpoints, with at least three annual observations per segment, adjacent joinpoints at least three years apart, and every segment (including the first and last) spanning at least three years; the span rule binds only for segments starting or ending within three years of the series bounds, and is stricter than the observation-count rule there; intervals use n − (2k + 2) residual degrees of freedom. APC = annual percent change within a segment (t-based 95% CI on the log scale); AAPC = span-weighted average APC (delta-method 95% CI). The AAPC is the span-weighted average of fitted segment slopes, not the endpoint-to-endpoint rate; the observed endpoints imply +2.49% per year for the Sub-Saharan Africa/High-income ratio. EFR = episode-fatality ratio (deaths per incident episode); gap = country EFR/within-super-region 10th-percentile frontier EFR. Episode denominators for intermediate years are linearly interpolated between the observed 1990, 2019 and 2023 values. Within each EFR segment the log-scale change decomposes additively into an observed-deaths component and an interpolated-episodes component; the denominator accounts for 42.5% (1990–2010), 61.7% (2010–2017) and 14.5% (2017–2023) of the absolute log movement (43.1%, 63.1%, 15.7% and 55.3% across the four segments of the previous three-joinpoint EFR fit, which is retained together with the previous median-gap fit in the supplementary joinpoint CSV). Under log-linear interpolation of the episode anchors the cumulative 2011–2017 EFR rise shrinks from +9.7% to +4.4% and is no longer monotone (+1.5% when the log-linear interpolation is applied country by country before aggregation).

### The Sub-Saharan Africa frontier problem

Within-region benchmarking understates the distance for regions whose frontier has stagnated. In 1990 the Sub-Saharan Africa frontier was defined by Cabo Verde, Uganda, South Africa, Eswatini and Kenya, whose EFRs (6.7–9.4 per 1,000 episodes) were already 2.9–4.0 times the High-income frontier of the same year. (With 46 countries in the region, the 10th-percentile frontier is interpolated between the fifth– and sixth-lowest country EFRs, so the 1990 frontier value, 9.85 per 1,000, sits just above this five-country group.) The region’s frontier improved from 9.85 to 6.93 per 1,000 between 1990 and 2023, about 30% in 33 years, while the High-income frontier improved by about two-thirds (2.32 to 0.73 per 1,000); the ratio of the two frontiers therefore widened from 4.2 to 9.5. Within the region, the number of countries at or near the frontier (gap ≤ 1.5) fell from 20 of 46 in 1990 to 13 in 2019, recovering only to 17 in 2023. Measured against the global 10th-percentile frontier of 2023, the median Sub-Saharan African country’s gap is 11.0, against 1.78 to its own regional frontier: within-region benchmarking reports less than one-sixth of the region’s true distance from achievable survival. The frontier ratio series is shown in Figure 5.

**Figure 3.**
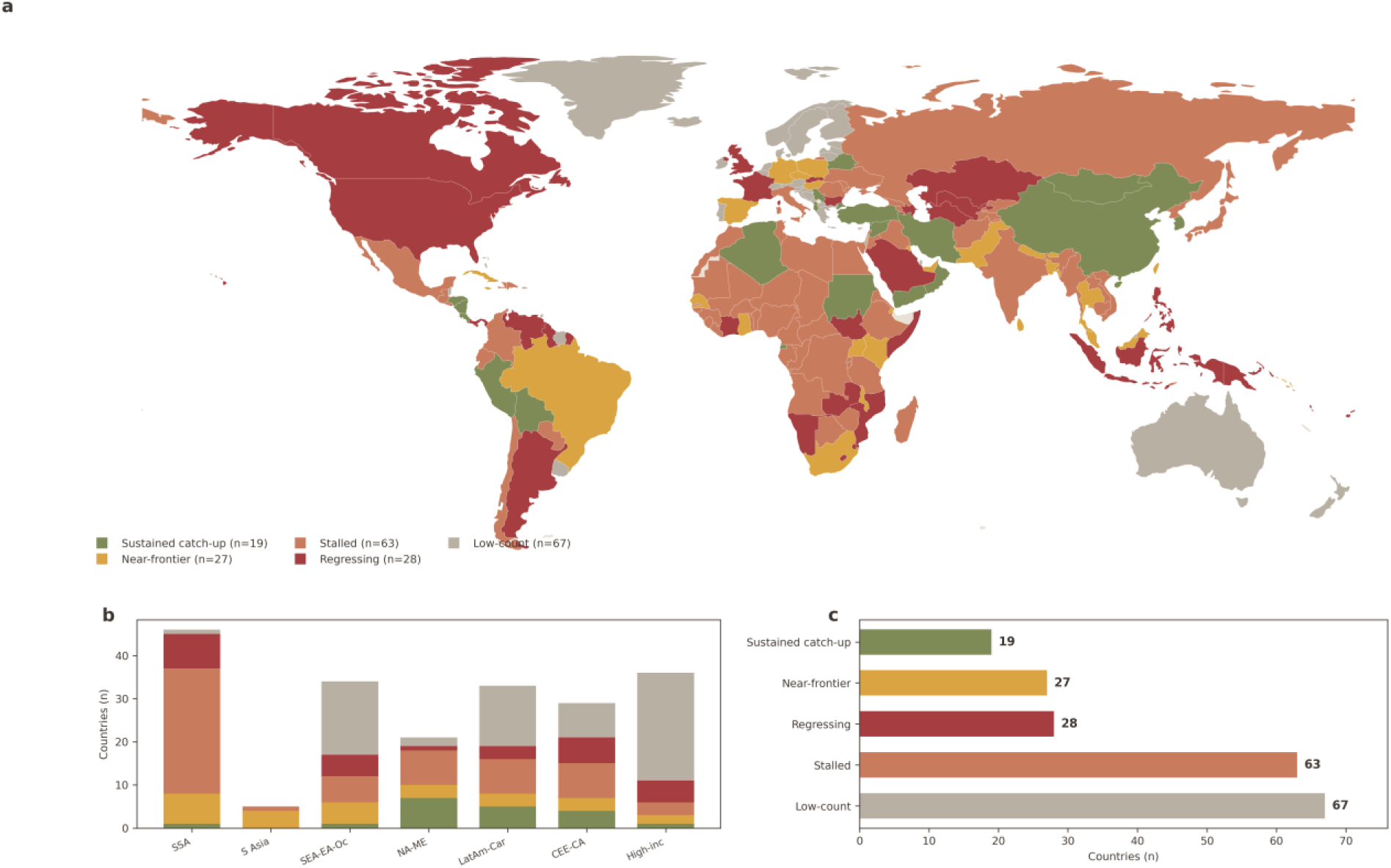
Catch-up phenotypes of 1990–2023 LRI EFR gap trajectories (anchors 1990, 2010, 2019, 2023). (a) Phenotype map: sustained catch-up (gap fell ≥30%; n=19), near-frontier (gap ≤1.5 throughout; n=27), stalled (n=63), regressing (gap rose ≥30%; n=28), low-count (<25 deaths in 2023; n=67). (b) Phenotype composition by GBD super-region (stacked country counts). (c) Phenotype counts: stalled 63, regressing 28, near-frontier 27, sustained catch-up 19, low-count 67.

**Figure 4.**
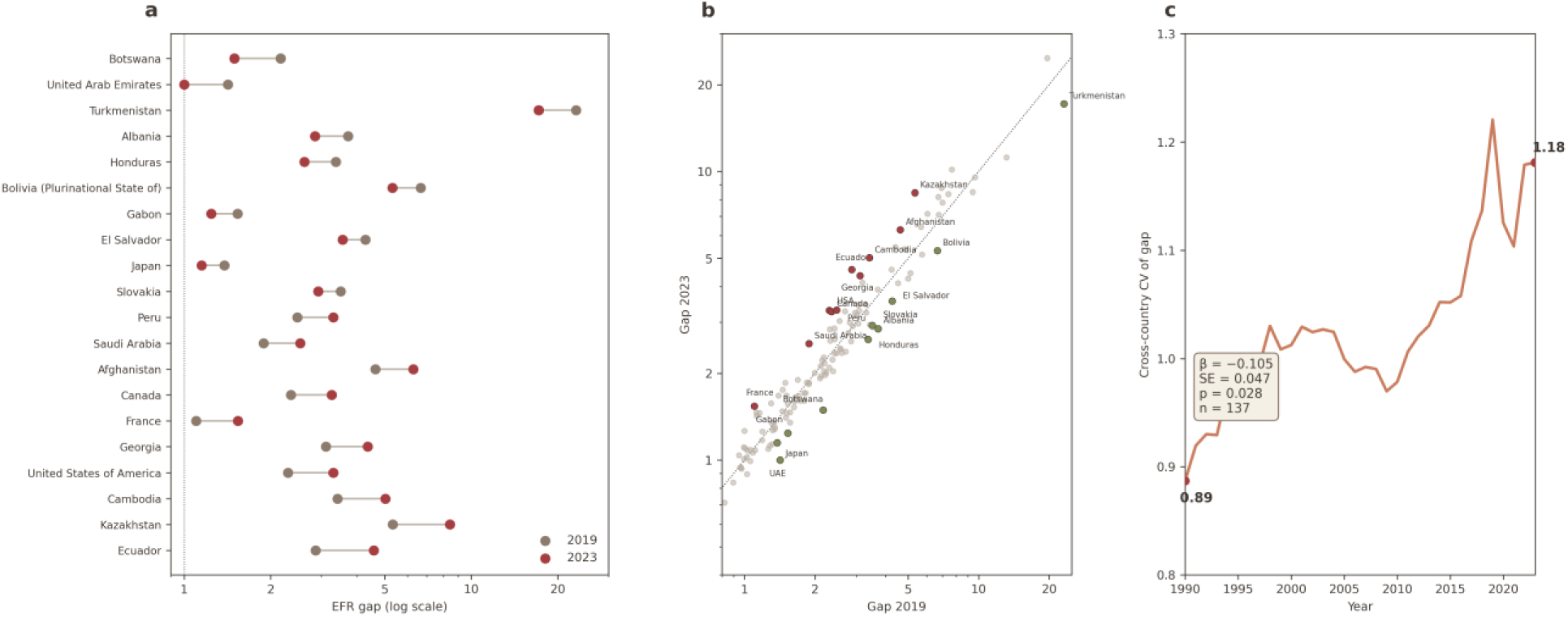
Gap change over the COVID-19 window, 2019 versus 2023 (low-count countries excluded). (a) Dumbbell plot of the ten fastest catch-up and ten largest regressing countries (log scale; grey dot = 2019, red dot = 2023). (b) Gap 2019 versus gap 2023 for the 137 eligible countries (log–log; dotted diagonal = no change); the 20 window movers are annotated. (c) Sigma/beta convergence statistics: cross-country coefficient of variation of the gap, 0.89 (1990) to 1.18 (2023); beta-convergence regression log(gap2023/gap1990) ∼ log(gap1990): slope β = −0.105 (SE 0.047, p = 0.028, n = 137).

**Figure 5.**
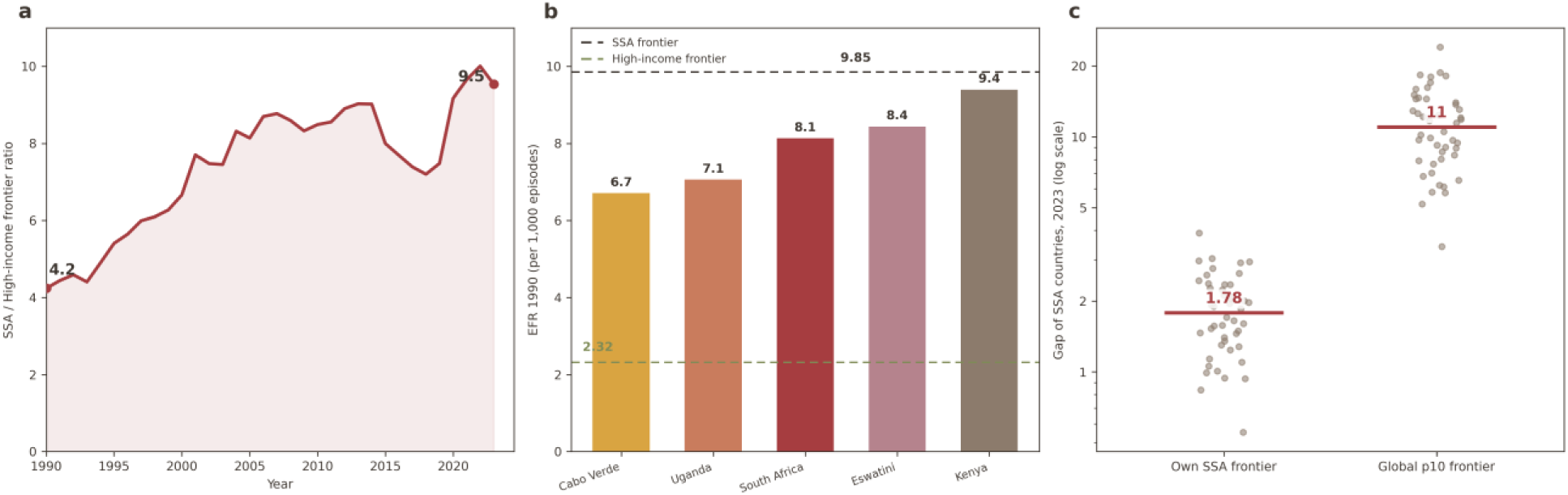
Sub-Saharan Africa frontier regression, 1990–2023. (a) Ratio of the SSA frontier EFR to the High-income frontier EFR by year: 4.2 (1990) to 9.5 (2023). (b) EFR in 1990 (per 1,000 episodes) of the five countries defining the SSA frontier — Cabo Verde 6.7, Uganda 7.1, South Africa 8.1, Eswatini 8.4, Kenya 9.4 — against the SSA frontier (9.85) and the High-income frontier (2.32). (c) Gap of the 46 SSA countries in 2023 to their own regional frontier (median 1.78) versus the global p10 frontier (median 11.0; log scale; horizontal bars = medians).

### URI cross-check and sensitivity analyses

The stalled-catch-up pattern is not specific to LRI. For URI, a near-zero-fatality contrast assembled without interpolation, the median gap was 4.15 in 1990, 4.36 in 2019 and 4.60 in 2023: no convergence over the same three decades, and 93.5% of avoidable URI deaths in 2023 occurred in Sub-Saharan Africa. Headline LRI patterns are qualitatively unchanged under 5th– and 25th-percentile frontiers (Table 5). The two external checks are likewise reassuring (Supplementary Table S7). National mean elevation correlated only weakly with the 2023 gap (eligible countries: Spearman ρ = 0.14, p = 0.11, n = 135; all countries with data: ρ = 0.19, p = 0.01, n = 174), so the wide gaps of high-altitude countries such as Bolivia, Mexico, Ecuador, Afghanistan, the Central Asian republics and Ethiopia are not explained by elevation alone. The UHC service coverage index correlated with the gap in the expected negative direction, but weakly (all countries with data, ρ = −0.18, p = 0.01, n = 193; eligible, ρ = −0.07, p = 0.42, n = 136), the attenuation expected, since within-region frontiers themselves track system quality.

## Discussion

### Principal findings

Across 204 countries and 33 years, the typical country made no net progress toward the best survival performance achieved within its own region. The median gap to the within-super-region LRI survival frontier was 1.86 in both 1990 and 2023, the share of countries more than twice their frontier rose by 2.0 percentage points (44.6% to 46.6%), and eligible countries split almost evenly between narrowing (67) and widening (69), with one unchanged. Sustained catch-up is real but rare and geographically concentrated: 19 countries, most of them in North Africa and the Middle East and Latin America, with China closing beyond its frontier entirely. Regression is similarly patterned, led by Central Asia but extending to high-income countries, including the United States. The pandemic window produced a sharp, only partly reversed divergence (median gap +10.8% by 2021, residual +3.1% by 2023). Where gaps meet burden, two distinct policy problems emerge: system-wide quality shortfalls in high-burden giants, and extreme outliers whose fatality per episode defies regional comparison. Finally, the region with the largest burden, Sub-Saharan Africa, is measured against a frontier that has itself fallen further behind global best practice, so within-region benchmarking reports a small fraction of the region’s true distance. Joinpoint analysis quantifies this verdict: the median gap’s 33-year average annual change (+0.03% per year, 95% CI −0.05 to 0.12) is statistically indistinguishable from zero, and the apparent mid-period rise in the global EFR is an artefact of the interpolated episode denominators rather than a reversal of survival improvement (Results; Table 6 note).

### Convergence is not the default trajectory

These results amount to a national-level falsification of the convergence assumption embedded in SDG-era child-survival discourse.^9^ Aggregate mortality fell 65% over the study period, a decline routinely read as evidence that lagging countries are catching up. The gap series shows the two processes are independent: deaths fell overwhelmingly because fewer children fell ill and because fatality improved almost everywhere, while relative positions against the moving frontier barely changed. Progress and convergence are not the same thing. This distinction matters for monitoring: a country can halve its LRI mortality while falling further behind its peers, and standard mortality tracking would record only success. The even split between narrowing and widening countries indicates that the diffusion mechanism assumed by convergence narratives has been weak for case survival in a cause group responsive to some of the oldest and cheapest interventions in child survival.^4^ The finding is consistent with uneven, development-decoupled gains in the HAQ Index^8^ and with the High Quality Health Systems Commission’s argument that the binding constraint has shifted from contact with services to the quality of care delivered.^7^ Between-country gaps in amenable mortality did narrow within Europe over recent decades;^6^ our results show that this regional experience does not generalize to a global, cause-specific case-survival metric.

### Two policy problems, not one

The priority grid separates countries for whom catch-up means different things. In the high-burden, moderate-gap giants (Nigeria, India, Niger, the Democratic Republic of the Congo), most avoidable deaths arise from average-quality case management applied to very large episode volumes; the relevant interventions are system-wide: reliable oxygen systems and pulse oximetry, timely effective antimicrobials, referral pathways for severe pneumonia, and sustained immunization against respiratory pathogens.^4,16^ A prospective Lives Saved Tool analysis independently corroborates this scale of opportunity: scaling four pneumonia interventions to at least 90% coverage in three high-burden countries would avert 45–58% of expected under-five pneumonia deaths,^17^ matching the avoidable shares implied here. The extreme-gap outliers present a different problem. A gap of 28.6 (Uzbekistan), 24.7 (Azerbaijan) or 17.2 (Turkmenistan) means children die once ill at rates an order of magnitude above the best decile of the same region; values of this size plausibly reflect case-management performance or data quality, including vital-registration misclassification; the appropriate first response is targeted clinical and data audit rather than inference from the point estimate alone. We deliberately refrain from adjudicating individual countries: the gap locates where scrutiny and support would yield the most, it does not assign cause.

### The anchoring effect of regional benchmarks

The Sub-Saharan Africa results expose a measurement trap with consequences beyond this study. Within-region benchmarking is designed to be fair and conservative, since a region’s best decile is an attained rather than aspirational standard; but when a region’s frontier stagnates, the benchmark anchors perception: the median Sub-Saharan African country appears 1.78 times from best practice against its own frontier while standing 11.0 times from the global frontier. The region’s frontier was already 4.2 times the High-income frontier in 1990 and is 9.5 times today, and the number of regional countries at or near their own frontier has shrunk. Two implications follow. First, regional frontier metrics should be reported alongside a global-frontier contrast, otherwise regions most in need of frontier-advancing investment will appear closest to target. Second, the binding constraint for Sub-Saharan Africa is not only laggard catch-up: even perfect within-region convergence would leave the region’s children dying once ill at about six times (6.2-fold) the rate the global best decile achieves. Closing that distance requires raising the frontier itself, through the quality-of-care investments the HQSS Commission set out,^7^ after which a functioning convergence mechanism, currently absent, would have something to propagate.

### High-income regression and the surveillance dialogue

The regression of high-income countries complicates any simple income gradient in catch-up. The United States doubled its gap over 33 years and widened it by a further 44% over the pandemic window; Canada and France show the same direction. These movements coincide with documented post-pandemic disruption and asynchronous resurgence of respiratory infections in high-income settings,^18^ and they demonstrate that frontier proximity is not a stable property of wealthy health systems but a performance variable that can deteriorate. The contrast with China is instructive for surveillance: one country moved from 2.4 times its regional frontier to below it, while another moved from 1.6 times to 3.3 times, and neither movement is visible in aggregate mortality trends, which improved in both. Annual gap tracking of the kind presented here is computable from standard GBD outputs at negligible cost and would make such reversals visible as they happen rather than in retrospect.

### Limitations

Seven limitations qualify these findings. First, the EFR is not a clinical case-fatality ratio: episode denominators include mild community illness, and the two are not interchangeable.^2^ Second, all inputs are GBD 2023 modelled estimates rather than counts; national gaps inherit the uncertainty of the underlying models, which is widest where vital registration is absent, and we report deterministic central values without propagated national uncertainty intervals. Third, LRI episodes for non-anchor years were linearly interpolated between the observed 1990, 2019 and 2023 anchors. The gap is exactly invariant to any rescaling applied uniformly to all countries within a super-region-year, but between-anchor trajectories are sensitive to the functional form of the interpolation: under log-linear interpolation the 2014 median gap is 1.74 rather than 1.86 and the median-gap joinpoints shift to 2012, 2016 and 2020. The exposure is greatest in the COVID-19 window: the 2021 median-gap peak (+10.8% versus 2019) rests on denominators interpolated across a period when respiratory incidence collapsed and rebounded non-linearly,^18^ so if true 2020–2021 incidence was lower than the interpolated path, the peak is understated; endpoint comparisons between observed years (2019 versus 2023) are unaffected. Fourth, ratio instability at low death counts led us to exclude 67 countries with fewer than 25 deaths in 2023 from classification; conclusions therefore concern the 137 countries where the burden is concentrated. The threshold is anchored to 2023 only: 30 of the excluded countries—the largest group high-income (12 of 30)—had counts above 25 in 1990, so their long-run trajectories are set aside even where the 1990 endpoint was precisely measured. Fifth, extreme gaps in Central Asia may partly reflect data-model artefacts, and we flag them for audit rather than interpret them literally. Sixth, the EFR is not age-standardized across the 0–19 band and reflects severity mix as well as care quality; a sensitivity analysis of this pipeline restricted to the under-five age band, which removes cross-country differences in child age structure by construction, left country avoidable-death rankings essentially unchanged (Spearman rank correlation 0.95 for LRI), so age composition is unlikely to explain the patterns reported. Seventh, gaps are not adjusted for elevation: hypobaric hypoxia at altitude may inflate measured episode fatality in high-altitude countries, so residual confounding would overstate, not understate, their gaps, although the weak elevation–gap correlation reported above bounds the likely bias. Phenotype thresholds are conventional; publishing the full country panel allows alternative classifications to be derived.

### Conclusions

For three decades, convergence toward the best survival performance within regions has been assumed but never tested at national level for a leading treatable cause of child death. Tested here across 204 countries, it did not occur: the median country ended 2023 exactly as far from its regional frontier as in 1990, nearly half of all countries remain more than twice their frontier, and widening countries slightly outnumber narrowing ones. Catch-up, where it happened, was concentrated and identifiable, in North Africa and the Middle East, parts of Latin America and China; regression was equally identifiable, led by Central Asia and extending to high-income settings. The pandemic window briefly widened the typical gap by a tenth, and that widening has only partly reversed. The residual burden of avoidable death is addressable through two distinct instruments: system-wide quality improvement in a small set of high-burden countries, and targeted audit where gaps are so extreme that case-management performance or data quality must be established first. And the Sub-Saharan Africa diagnostic shows that the benchmark itself can deceive: a region measured against a stagnating frontier will always look closer to best practice than it is. The pattern is not an artefact of one cause group: for upper respiratory infections, a near-zero-fatality contrast observed without interpolation, the median gap widened over the same three decades. Three implications follow for the SDG era’s monitoring architecture. Aggregate mortality trends should be read alongside episode-based survival metrics, since the first can improve while the second stagnates. Regional benchmarks should be reported with a global-frontier contrast, so that frontier stagnation cannot masquerade as proximity to best practice. And convergence itself should be treated as an outcome to be measured, not a tendency to be assumed. Convergence, if it is wanted, will have to be built: tracked annually, country by country, in the metric that separates falling ill from dying once ill.

## Declarations

### Contributors

CS conceived and designed the study. CS developed the analytical framework. CS, DL and HC curated the data and performed the analyses. XW prepared the figures and assembled the supplementary materials. CS drafted the manuscript. DL, HC, and CS interpreted the results and critically revised the manuscript. CS supervised the work. All authors had full access to all of the data in the study, verified the underlying data, accept responsibility for the decision to submit for publication, and read and approved the final version.

### Presentation

This work has not been presented at any scientific meeting.

### Data sharing

All inputs are publicly available from the Global Burden of Disease Study 2023 results tools. The analytic country-year panel, phenotype classifications and all derived tables accompany this article (Supplementary Tables S1–S7). Version 3 additionally provides the joinpoint analysis script (joinpoint_efr_trajectories.py), its input panel (lri_gap_panel_country_year.csv) and its outputs (Supplementary_Joinpoint_NoConvergenceEFR.csv, Supplementary_Joinpoint_series_NoConvergenceEFR.csv, and the previous-BIC comparison fits in Supplementary_Joinpoint_legacyBIC_NoConvergenceEFR.csv) in the accompanying code folder.

## Data Availability

All inputs are publicly available from the Global Burden of Disease Study 2023 results tools. The analytic country-year panel, phenotype classifications and all derived tables accompany this article (Supplementary Tables S1-S7). Version 3 additionally provides the joinpoint analysis script (joinpoint_efr_trajectories.py), its input panel (lri_gap_panel_country_year.csv) and its outputs (Supplementary_Joinpoint_NoConvergenceEFR.csv, Supplementary_Joinpoint_series_NoConvergenceEFR.csv, and the previous-BIC comparison fits in Supplementary_Joinpoint_legacyBIC_NoConvergenceEFR.csv) in the accompanying code folder. Reporting follows the GATHER statement.

https://vizhub.healthdata.org/gbd-results/

