## Supplementary_Materials for "No convergence in three decades: national trajectories of episode-fatality ratios for childhood lower respiratory infections in 204 countries, 1990-2023"

### Supplementary Tables S1–S7

#### Table S1. Country-level EFR gap anchors, 2023 burden and catch-up phenotypes for the 137 eligible countries

| Country | ISO3 | Super-region | Gap 1990 | Gap 2010 | Gap 2019 | Gap 2023 | LRI deaths 2023 | Avoidable deaths 2023 | EFR 2023 (per 1,000 episodes) | Phenotype | COVID ratio | Long-run ratio |
|---|---|---|---|---|---|---|---|---|---|---|---|---|
| Afghanistan | AFG | North Africa and Middle East | 6.00 | 6.90 | 4.64 | 6.29 | 10,268 | 8,634 | 10.06 | Stalled | 1.36 | 1.05 |
| Albania | ALB | Central Europe, Eastern Europe, and Central Asia | 7.13 | 2.56 | 3.72 | 2.86 | 28 | 18 | 3.36 | Sustained catch-up | 0.77 | 0.40 |
| Algeria | DZA | North Africa and Middle East | 2.46 | 1.98 | 1.70 | 1.67 | 1,137 | 455 | 2.67 | Sustained catch-up | 0.98 | 0.68 |
| Angola | AGO | Sub-Saharan Africa | 2.40 | 2.53 | 2.47 | 2.35 | 10,277 | 5,905 | 16.28 | Stalled | 0.95 | 0.98 |
| Argentina | ARG | High-income | 2.88 | 5.44 | 4.25 | 4.57 | 436 | 340 | 3.32 | Regressing | 1.08 | 1.59 |
| Armenia | ARM | Central Europe, Eastern Europe, and Central Asia | 7.24 | 6.07 | 7.41 | 8.35 | 146 | 128 | 9.82 | Stalled | 1.13 | 1.15 |
| Azerbaijan | AZE | Central Europe, Eastern Europe, and Central Asia | 11.37 | 13.74 | 19.66 | 24.72 | 1,964 | 1,884 | 29.08 | Regressing | 1.26 | 2.17 |
| Bangladesh | BGD | South Asia | 1.09 | 0.94 | 1.06 | 1.08 | 14,891 | 1,159 | 6.09 | Near-frontier | 1.02 | 0.99 |

| Country | ISO3 | Super-region | Gap 1990 | Gap 2010 | Gap 2019 | Gap 2023 | LRI deaths 2023 | Avoidable deaths 2023 | EFR 2023 (per 1,000 episodes) | Phenotype | COVID ratio | Long-run ratio |
|---|---|---|---|---|---|---|---|---|---|---|---|---|
| Belarus | BLR | Central Europe, Eastern Europe, and Central Asia | 1.80 | 1.00 | 0.82 | 0.71 | 28 | 0 | 0.84 | Sustained catch-up | 0.87 | 0.40 |
| Benin | BEN | Sub-Saharan Africa | 2.48 | 2.08 | 2.24 | 2.01 | 4,388 | 2,201 | 13.90 | Stalled | 0.90 | 0.81 |
| Bhutan | BTN | South Asia | 1.06 | 1.10 | 1.31 | 1.35 | 41 | 11 | 7.57 | Near-frontier | 1.03 | 1.27 |
| Bolivia (Plurinational State of) | BOL | Latin America and Caribbean | 11.10 | 9.84 | 6.66 | 5.32 | 1,388 | 1,127 | 11.32 | Sustained catch-up | 0.80 | 0.48 |
| Botswana | BWA | Sub-Saharan Africa | 1.20 | 2.11 | 2.17 | 1.49 | 295 | 98 | 10.34 | Stalled | 0.69 | 1.25 |
| Brazil | BRA | Latin America and Caribbean | 0.93 | 0.76 | 1.03 | 1.08 | 3,841 | 285 | 2.30 | Near-frontier | 1.05 | 1.16 |
| Bulgaria | BGR | Central Europe, Eastern Europe, and Central Asia | 2.13 | 2.75 | 3.29 | 3.47 | 83 | 59 | 4.08 | Regressing | 1.05 | 1.63 |
| Burkina Faso | BFA | Sub-Saharan Africa | 2.64 | 1.83 | 2.18 | 2.27 | 7,750 | 4,337 | 15.73 | Stalled | 1.04 | 0.86 |
| Burundi | BDI | Sub-Saharan Africa | 1.74 | 1.57 | 1.69 | 1.90 | 5,659 | 2,677 | 13.15 | Stalled | 1.12 | 1.09 |
| Cambodia | KHM | Southeast Asia, East Asia, and Oceania | 4.79 | 4.07 | 3.42 | 5.02 | 2,185 | 1,750 | 10.07 | Stalled | 1.47 | 1.05 |
| Cameroon | CMR | Sub-Saharan Africa | 2.13 | 2.60 | 2.57 | 2.44 | 10,533 | 6,219 | 16.91 | Stalled | 0.95 | 1.15 |
| Canada | CAN | High-income | 1.90 | 2.28 | 2.35 | 3.27 | 50 | 35 | 2.37 | Regressing | 1.39 | 1.72 |
| Central African Republic | CAF | Sub-Saharan Africa | 2.35 | 2.41 | 2.90 | 2.91 | 4,948 | 3,249 | 20.18 | Stalled | 1.01 | 1.24 |

| Country | ISO3 | Super-region | Gap 1990 | Gap 2010 | Gap 2019 | Gap 2023 | LRI deaths 2023 | Avoidable deaths 2023 | EFR 2023 (per 1,000 episodes) | Phenotype | COVID ratio | Long-run ratio |
|---|---|---|---|---|---|---|---|---|---|---|---|---|
| Chad | TCD | Sub-Saharan Africa | 2.22 | 2.09 | 2.43 | 2.63 | 13,920 | 8,621 | 18.20 | Stalled | 1.08 | 1.18 |
| Chile | CHL | High-income | 4.07 | 2.63 | 2.69 | 3.28 | 85 | 59 | 2.38 | Stalled | 1.22 | 0.81 |
| China | CHN | Southeast Asia, East Asia, and Oceania | 2.43 | 0.84 | 0.50 | 0.50 | 7,423 | 0 | 0.99 | Sustained catch-up | 0.98 | 0.20 |
| Colombia | COL | Latin America and Caribbean | 3.23 | 3.54 | 2.82 | 2.99 | 1,108 | 738 | 6.37 | Stalled | 1.06 | 0.93 |
| Comoros | COM | Sub-Saharan Africa | 1.23 | 0.90 | 1.30 | 1.58 | 186 | 68 | 10.91 | Stalled | 1.21 | 1.28 |
| Congo | COG | Sub-Saharan Africa | 1.39 | 1.68 | 1.53 | 1.57 | 1,006 | 365 | 10.86 | Stalled | 1.03 | 1.13 |
| Costa Rica | CRI | Latin America and Caribbean | 2.86 | 1.80 | 1.88 | 1.84 | 45 | 20 | 3.91 | Sustained catch-up | 0.97 | 0.64 |
| Cuba | CUB | Latin America and Caribbean | 1.27 | 1.08 | 1.16 | 1.46 | 109 | 34 | 3.10 | Near-frontier | 1.26 | 1.15 |
| Czechia | CZE | Central Europe, Eastern Europe, and Central Asia | 1.15 | 1.04 | 1.45 | 1.48 | 53 | 17 | 1.74 | Near-frontier | 1.02 | 1.28 |
| Côte d'Ivoire | CIV | Sub-Saharan Africa | 2.06 | 2.05 | 2.77 | 2.76 | 11,371 | 7,246 | 19.10 | Regressing | 0.99 | 1.34 |
| Democratic People's Republic of Korea | PRK | Southeast Asia, East Asia, and Oceania | 1.39 | 1.85 | 1.45 | 1.75 | 745 | 320 | 3.51 | Stalled | 1.21 | 1.26 |
| Democratic Republic of the Congo | COD | Sub-Saharan Africa | 1.68 | 1.92 | 2.10 | 1.92 | 30,177 | 14,470 | 13.31 | Stalled | 0.92 | 1.15 |

| Country | ISO3 | Super-region | Gap 1990 | Gap 2010 | Gap 2019 | Gap 2023 | LRI deaths 2023 | Avoidable deaths 2023 | EFR 2023 (per 1,000 episodes) | Phenotype | COVID ratio | Long-run ratio |
|---|---|---|---|---|---|---|---|---|---|---|---|---|
| Djibouti | DJI | Sub-Saharan Africa | 1.06 | 1.34 | 1.34 | 1.30 | 231 | 54 | 9.03 | Near-frontier | 0.97 | 1.23 |
| Dominican Republic | DOM | Latin America and Caribbean | 2.54 | 2.90 | 2.57 | 2.43 | 323 | 190 | 5.17 | Stalled | 0.94 | 0.96 |
| Ecuador | ECU | Latin America and Caribbean | 3.94 | 4.41 | 2.87 | 4.57 | 1,210 | 945 | 9.74 | Stalled | 1.59 | 1.16 |
| Egypt | EGY | North Africa and Middle East | 5.06 | 4.23 | 3.19 | 4.13 | 10,096 | 7,649 | 6.60 | Stalled | 1.29 | 0.82 |
| El Salvador | SLV | Latin America and Caribbean | 4.92 | 4.82 | 4.28 | 3.56 | 177 | 127 | 7.58 | Stalled | 0.83 | 0.72 |
| Equatorial Guinea | GNQ | Sub-Saharan Africa | 2.29 | 1.41 | 1.50 | 1.40 | 209 | 60 | 9.70 | Sustained catch-up | 0.93 | 0.61 |
| Eritrea | ERI | Sub-Saharan Africa | 1.34 | 1.44 | 1.82 | 1.71 | 2,420 | 1,002 | 11.82 | Stalled | 0.94 | 1.27 |
| Eswatini | SWZ | Sub-Saharan Africa | 0.86 | 2.75 | 3.43 | 2.94 | 327 | 216 | 20.37 | Regressing | 0.86 | 3.43 |
| Ethiopia | ETH | Sub-Saharan Africa | 1.58 | 1.17 | 1.35 | 1.28 | 21,197 | 4,672 | 8.89 | Stalled | 0.95 | 0.81 |
| Fiji | FJI | Southeast Asia, East Asia, and Oceania | 1.45 | 2.78 | 2.32 | 2.84 | 61 | 40 | 5.69 | Regressing | 1.23 | 1.95 |
| France | FRA | High-income | 1.02 | 1.05 | 1.10 | 1.54 | 70 | 24 | 1.12 | Regressing | 1.40 | 1.51 |
| Gabon | GAB | Sub-Saharan Africa | 1.11 | 1.19 | 1.53 | 1.24 | 209 | 40 | 8.59 | Stalled | 0.81 | 1.12 |
| Gambia | GMB | Sub-Saharan Africa | 1.55 | 1.62 | 2.21 | 1.97 | 733 | 361 | 13.65 | Stalled | 0.89 | 1.27 |

| Country | ISO3 | Super-region | Gap 1990 | Gap 2010 | Gap 2019 | Gap 2023 | LRI deaths 2023 | Avoidable deaths 2023 | EFR 2023 (per 1,000 episodes) | Phenotype | COVID ratio | Long-run ratio |
|---|---|---|---|---|---|---|---|---|---|---|---|---|
| Georgia | GEO | Central Europe, Eastern Europe, and Central Asia | 5.50 | 2.72 | 3.11 | 4.36 | 92 | 71 | 5.12 | Stalled | 1.40 | 0.79 |
| Germany | DEU | High-income | 1.14 | 0.96 | 1.02 | 0.90 | 45 | 0 | 0.65 | Near-frontier | 0.88 | 0.79 |
| Ghana | GHA | Sub-Saharan Africa | 1.32 | 1.05 | 1.32 | 1.14 | 3,929 | 469 | 7.87 | Near-frontier | 0.86 | 0.86 |
| Guatemala | GTM | Latin America and Caribbean | 10.32 | 10.96 | 7.67 | 10.16 | 2,312 | 2,084 | 21.63 | Stalled | 1.32 | 0.98 |
| Guinea | GIN | Sub-Saharan Africa | 3.15 | 2.44 | 2.85 | 2.58 | 7,170 | 4,392 | 17.88 | Stalled | 0.90 | 0.82 |
| Guinea-Bissau | GNB | Sub-Saharan Africa | 2.13 | 2.03 | 2.13 | 1.96 | 650 | 318 | 13.55 | Stalled | 0.92 | 0.92 |
| Guyana | GUY | Latin America and Caribbean | 3.67 | 4.38 | 5.72 | 5.15 | 45 | 36 | 10.98 | Regressing | 0.90 | 1.40 |
| Haiti | HTI | Latin America and Caribbean | 10.49 | 12.06 | 9.66 | 9.55 | 3,774 | 3,379 | 20.35 | Stalled | 0.99 | 0.91 |
| Honduras | HND | Latin America and Caribbean | 4.98 | 4.53 | 3.37 | 2.62 | 288 | 178 | 5.58 | Sustained catch-up | 0.78 | 0.53 |
| Hungary | HUN | Central Europe, Eastern Europe, and Central Asia | 0.81 | 0.57 | 0.78 | 0.81 | 27 | 0 | 0.95 | Near-frontier | 1.04 | 0.99 |
| India | IND | South Asia | 1.35 | 1.83 | 1.80 | 1.61 | 143,193 | 54,109 | 9.03 | Stalled | 0.89 | 1.19 |
| Indonesia | IDN | Southeast Asia, East Asia, and Oceania | 2.40 | 2.92 | 3.02 | 3.20 | 14,197 | 9,757 | 6.41 | Regressing | 1.06 | 1.33 |

| Country | ISO3 | Super-region | Gap 1990 | Gap 2010 | Gap 2019 | Gap 2023 | LRI deaths 2023 | Avoidable deaths 2023 | EFR 2023 (per 1,000 episodes) | Phenotype | COVID ratio | Long-run ratio |
|---|---|---|---|---|---|---|---|---|---|---|---|---|
| Iran (Islamic Republic of) | IRN | North Africa and Middle East | 2.63 | 1.61 | 1.51 | 1.68 | 1,058 | 426 | 2.68 | Sustained catch-up | 1.11 | 0.64 |
| Iraq | IRQ | North Africa and Middle East | 2.43 | 3.61 | 2.19 | 2.17 | 1,835 | 988 | 3.47 | Stalled | 0.99 | 0.89 |
| Italy | ITA | High-income | 1.59 | 1.19 | 1.49 | 1.85 | 44 | 20 | 1.35 | Stalled | 1.24 | 1.17 |
| Jamaica | JAM | Latin America and Caribbean | 1.42 | 1.10 | 1.11 | 1.08 | 26 | 2 | 2.30 | Near-frontier | 0.98 | 0.76 |
| Japan | JPN | High-income | 1.00 | 1.64 | 1.38 | 1.15 | 91 | 12 | 0.83 | Stalled | 0.83 | 1.15 |
| Jordan | JOR | North Africa and Middle East | 2.11 | 2.39 | 1.80 | 1.71 | 384 | 159 | 2.73 | Stalled | 0.95 | 0.81 |
| Kazakhstan | KAZ | Central Europe, Eastern Europe, and Central Asia | 5.98 | 4.94 | 5.33 | 8.44 | 1,212 | 1,068 | 9.93 | Regressing | 1.58 | 1.41 |
| Kenya | KEN | Sub-Saharan Africa | 0.95 | 1.31 | 0.90 | 0.84 | 8,348 | 0 | 5.80 | Near-frontier | 0.94 | 0.88 |
| Kuwait | KWT | North Africa and Middle East | 0.91 | 1.12 | 1.19 | 1.17 | 77 | 11 | 1.88 | Near-frontier | 0.99 | 1.29 |
| Kyrgyzstan | KGZ | Central Europe, Eastern Europe, and Central Asia | 8.37 | 8.25 | 7.00 | 7.82 | 538 | 469 | 9.20 | Stalled | 1.12 | 0.93 |
| Lao People's Democratic Republic | LAO | Southeast Asia, East Asia, and Oceania | 8.66 | 8.26 | 6.93 | 8.78 | 1,934 | 1,713 | 17.59 | Stalled | 1.27 | 1.01 |

| Country | ISO3 | Super-region | Gap 1990 | Gap 2010 | Gap 2019 | Gap 2023 | LRI deaths 2023 | Avoidable deaths 2023 | EFR 2023 (per 1,000 episodes) | Phenotype | COVID ratio | Long-run ratio |
|---|---|---|---|---|---|---|---|---|---|---|---|---|
| Lebanon | LBN | North Africa and Middle East | 4.29 | 2.90 | 3.31 | 3.25 | 252 | 175 | 5.20 | Stalled | 0.98 | 0.76 |
| Lesotho | LSO | Sub-Saharan Africa | 1.13 | 1.55 | 1.57 | 1.47 | 526 | 167 | 10.16 | Regressing | 0.94 | 1.30 |
| Liberia | LBR | Sub-Saharan Africa | 2.94 | 2.14 | 2.43 | 2.35 | 1,919 | 1,103 | 16.28 | Stalled | 0.97 | 0.80 |
| Libya | LBY | North Africa and Middle East | 1.95 | 1.50 | 1.39 | 1.67 | 182 | 73 | 2.68 | Stalled | 1.20 | 0.86 |
| Madagascar | MDG | Sub-Saharan Africa | 1.92 | 1.87 | 2.38 | 2.23 | 12,892 | 7,108 | 15.44 | Stalled | 0.94 | 1.16 |
| Malawi | MWI | Sub-Saharan Africa | 1.22 | 0.93 | 0.97 | 0.93 | 2,789 | 0 | 6.47 | Near-frontier | 0.96 | 0.77 |
| Malaysia | MYS | Southeast Asia, East Asia, and Oceania | 1.00 | 1.00 | 1.02 | 1.02 | 471 | 10 | 2.05 | Near-frontier | 1.00 | 1.03 |
| Mali | MLI | Sub-Saharan Africa | 2.47 | 1.90 | 2.39 | 2.04 | 7,788 | 3,964 | 14.11 | Stalled | 0.85 | 0.82 |
| Mauritania | MRT | Sub-Saharan Africa | 1.63 | 1.30 | 1.34 | 1.35 | 873 | 228 | 9.37 | Stalled | 1.01 | 0.83 |
| Mexico | MEX | Latin America and Caribbean | 9.02 | 6.77 | 6.73 | 8.17 | 3,810 | 3,344 | 17.41 | Stalled | 1.21 | 0.91 |
| Mongolia | MNG | Central Europe, Eastern Europe, and Central Asia | 13.00 | 10.57 | 9.42 | 8.48 | 277 | 244 | 9.97 | Sustained catch-up | 0.90 | 0.65 |
| Morocco | MAR | North Africa and Middle East | 3.29 | 3.35 | 2.61 | 2.35 | 1,100 | 633 | 3.76 | Stalled | 0.90 | 0.71 |
| Mozambique | MOZ | Sub-Saharan Africa | 1.34 | 1.67 | 2.12 | 2.12 | 9,097 | 4,808 | 14.69 | Regressing | 1.00 | 1.58 |

| Country | ISO3 | Super-region | Gap 1990 | Gap 2010 | Gap 2019 | Gap 2023 | LRI deaths 2023 | Avoidable deaths 2023 | EFR 2023 (per 1,000 episodes) | Phenotype | COVID ratio | Long-run ratio |
|---|---|---|---|---|---|---|---|---|---|---|---|---|
| Myanmar | MMR | Southeast Asia, East Asia, and Oceania | 5.51 | 5.93 | 5.67 | 6.43 | 8,296 | 7,006 | 12.89 | Stalled | 1.13 | 1.17 |
| Namibia | NAM | Sub-Saharan Africa | 1.05 | 1.46 | 1.63 | 1.53 | 520 | 179 | 10.56 | Regressing | 0.94 | 1.46 |
| Nepal | NPL | South Asia | 1.36 | 1.17 | 1.19 | 1.25 | 2,600 | 528 | 7.05 | Near-frontier | 1.05 | 0.92 |
| Nicaragua | NIC | Latin America and Caribbean | 7.06 | 6.49 | 4.53 | 4.11 | 266 | 202 | 8.75 | Sustained catch-up | 0.91 | 0.58 |
| Niger | NER | Sub-Saharan Africa | 3.28 | 2.79 | 3.71 | 3.90 | 25,191 | 18,738 | 27.04 | Stalled | 1.05 | 1.19 |
| Nigeria | NGA | Sub-Saharan Africa | 2.83 | 2.68 | 2.71 | 2.38 | 116,482 | 67,490 | 16.47 | Stalled | 0.88 | 0.84 |
| Oman | OMN | North Africa and Middle East | 2.16 | 0.97 | 1.26 | 1.09 | 74 | 6 | 1.75 | Sustained catch-up | 0.87 | 0.51 |
| Pakistan | PAK | South Asia | 0.96 | 1.12 | 0.96 | 0.94 | 36,134 | 0 | 5.30 | Near-frontier | 0.98 | 0.99 |
| Palestine | PSE | North Africa and Middle East | 1.32 | 1.41 | 1.00 | 1.26 | 161 | 34 | 2.02 | Near-frontier | 1.26 | 0.95 |
| Panama | PAN | Latin America and Caribbean | 3.89 | 7.15 | 4.42 | 5.47 | 170 | 139 | 11.64 | Regressing | 1.24 | 1.40 |
| Papua New Guinea | PNG | Southeast Asia, East Asia, and Oceania | 4.22 | 8.74 | 6.74 | 7.10 | 3,314 | 2,847 | 14.23 | Regressing | 1.05 | 1.68 |
| Paraguay | PRY | Latin America and Caribbean | 2.20 | 2.99 | 1.89 | 1.86 | 164 | 76 | 3.96 | Stalled | 0.98 | 0.85 |

| Country | ISO3 | Super-region | Gap 1990 | Gap 2010 | Gap 2019 | Gap 2023 | LRI deaths 2023 | Avoidable deaths 2023 | EFR 2023 (per 1,000 episodes) | Phenotype | COVID ratio | Long-run ratio |
|---|---|---|---|---|---|---|---|---|---|---|---|---|
| Peru | PER | Latin America and Caribbean | 5.80 | 3.01 | 2.48 | 3.31 | 1,972 | 1,376 | 7.05 | Sustained catch-up | 1.34 | 0.57 |
| Philippines | PHL | Southeast Asia, East Asia, and Oceania | 2.21 | 3.32 | 3.06 | 3.45 | 8,767 | 6,223 | 6.91 | Regressing | 1.13 | 1.56 |
| Poland | POL | Central Europe, Eastern Europe, and Central Asia | 0.96 | 0.73 | 0.72 | 0.86 | 156 | 0 | 1.01 | Near-frontier | 1.20 | 0.89 |
| Republic of Korea | KOR | High-income | 2.32 | 0.93 | 0.95 | 1.04 | 33 | 1 | 0.76 | Sustained catch-up | 1.10 | 0.45 |
| Republic of Moldova | MDA | Central Europe, Eastern Europe, and Central Asia | 3.82 | 4.49 | 5.00 | 4.26 | 56 | 43 | 5.01 | Stalled | 0.85 | 1.12 |
| Romania | ROU | Central Europe, Eastern Europe, and Central Asia | 4.28 | 4.45 | 5.10 | 4.44 | 326 | 253 | 5.23 | Stalled | 0.87 | 1.04 |
| Russian Federation | RUS | Central Europe, Eastern Europe, and Central Asia | 2.94 | 3.14 | 2.33 | 2.60 | 1,016 | 624 | 3.05 | Stalled | 1.12 | 0.88 |
| Rwanda | RWA | Sub-Saharan Africa | 1.66 | 1.31 | 1.65 | 1.65 | 3,302 | 1,305 | 11.45 | Stalled | 1.00 | 1.00 |
| Sao Tome and Principe | STP | Sub-Saharan Africa | 1.36 | 1.50 | 1.08 | 1.06 | 25 | 1 | 7.34 | Stalled | 0.98 | 0.78 |
| Saudi Arabia | SAU | North Africa and Middle East | 1.62 | 1.37 | 1.89 | 2.54 | 1,016 | 616 | 4.06 | Regressing | 1.34 | 1.56 |

| Country | ISO3 | Super-region | Gap 1990 | Gap 2010 | Gap 2019 | Gap 2023 | LRI deaths 2023 | Avoidable deaths 2023 | EFR 2023 (per 1,000 episodes) | Phenotype | COVID ratio | Long-run ratio |
|---|---|---|---|---|---|---|---|---|---|---|---|---|
| Senegal | SEN | Sub-Saharan Africa | 1.45 | 0.95 | 1.00 | 1.01 | 2,371 | 19 | 6.98 | Near-frontier | 1.01 | 0.69 |
| Serbia | SRB | Central Europe, Eastern Europe, and Central Asia | 1.96 | 1.03 | 1.56 | 1.35 | 29 | 7 | 1.59 | Sustained catch-up | 0.86 | 0.69 |
| Sierra Leone | SLE | Sub-Saharan Africa | 2.37 | 2.55 | 3.06 | 2.97 | 3,574 | 2,370 | 20.57 | Stalled | 0.97 | 1.25 |
| Slovakia | SVK | Central Europe, Eastern Europe, and Central Asia | 1.91 | 2.30 | 3.51 | 2.93 | 63 | 42 | 3.44 | Regressing | 0.83 | 1.53 |
| Solomon Islands | SLB | Southeast Asia, East Asia, and Oceania | 1.26 | 1.37 | 1.13 | 1.43 | 46 | 14 | 2.86 | Near-frontier | 1.27 | 1.13 |
| Somalia | SOM | Sub-Saharan Africa | 1.05 | 1.69 | 1.84 | 1.86 | 8,812 | 4,068 | 12.87 | Regressing | 1.01 | 1.77 |
| South Africa | ZAF | Sub-Saharan Africa | 0.83 | 1.38 | 1.06 | 0.99 | 6,418 | 0 | 6.87 | Near-frontier | 0.94 | 1.20 |
| South Sudan | SSD | Sub-Saharan Africa | 2.08 | 2.02 | 2.55 | 3.04 | 4,962 | 3,328 | 21.04 | Regressing | 1.19 | 1.46 |
| Spain | ESP | High-income | 1.33 | 1.35 | 1.12 | 1.46 | 34 | 11 | 1.06 | Near-frontier | 1.30 | 1.10 |
| Sri Lanka | LKA | Southeast Asia, East Asia, and Oceania | 0.74 | 0.79 | 0.60 | 0.70 | 197 | 0 | 1.40 | Near-frontier | 1.16 | 0.94 |
| Sudan | SDN | North Africa and Middle East | 6.72 | 6.03 | 3.09 | 3.29 | 2,591 | 1,805 | 5.27 | Sustained catch-up | 1.06 | 0.49 |
| Syrian Arab Republic | SYR | North Africa and Middle East | 1.83 | 3.03 | 1.32 | 1.27 | 380 | 81 | 2.03 | Sustained catch-up | 0.96 | 0.70 |

| Country | ISO3 | Super-region | Gap 1990 | Gap 2010 | Gap 2019 | Gap 2023 | LRI deaths 2023 | Avoidable deaths 2023 | EFR 2023 (per 1,000 episodes) | Phenotype | COVID ratio | Long-run ratio |
|---|---|---|---|---|---|---|---|---|---|---|---|---|
| Taiwan | TWN | Southeast Asia, East Asia, and Oceania | 0.41 | 0.30 | 0.30 | 0.38 | 79 | 0 | 0.76 | Near-frontier | 1.27 | 0.93 |
| Tajikistan | TJK | Central Europe, Eastern Europe, and Central Asia | 9.89 | 14.54 | 13.17 | 11.20 | 1,635 | 1,489 | 13.17 | Stalled | 0.85 | 1.13 |
| Thailand | THA | Southeast Asia, East Asia, and Oceania | 0.77 | 1.01 | 0.99 | 1.11 | 682 | 66 | 2.22 | Near-frontier | 1.12 | 1.43 |
| Timor-Leste | TLS | Southeast Asia, East Asia, and Oceania | 6.06 | 4.47 | 4.82 | 5.39 | 342 | 278 | 10.79 | Stalled | 1.12 | 0.89 |
| Togo | TGO | Sub-Saharan Africa | 1.67 | 1.67 | 1.74 | 1.61 | 1,940 | 732 | 11.12 | Stalled | 0.92 | 0.96 |
| Tunisia | TUN | North Africa and Middle East | 2.36 | 2.30 | 1.99 | 2.01 | 304 | 153 | 3.22 | Stalled | 1.01 | 0.85 |
| Turkmenistan | TKM | Central Europe, Eastern Europe, and Central Asia | 9.46 | 14.35 | 23.13 | 17.17 | 1,192 | 1,122 | 20.20 | Regressing | 0.74 | 1.81 |
| Türkiye | TUR | North Africa and Middle East | 3.41 | 1.07 | 1.28 | 1.12 | 947 | 103 | 1.80 | Sustained catch-up | 0.87 | 0.33 |
| Uganda | UGA | Sub-Saharan Africa | 0.72 | 0.86 | 0.96 | 0.94 | 6,676 | 0 | 6.53 | Near-frontier | 0.98 | 1.31 |
| Ukraine | UKR | Central Europe, Eastern Europe, and Central Asia | 1.93 | 1.73 | 2.19 | 2.03 | 184 | 93 | 2.39 | Stalled | 0.93 | 1.05 |

| Country | ISO3 | Super-region | Gap 1990 | Gap 2010 | Gap 2019 | Gap 2023 | LRI deaths 2023 | Avoidable deaths 2023 | EFR 2023 (per 1,000 episodes) | Phenotype | COVID ratio | Long-run ratio |
|---|---|---|---|---|---|---|---|---|---|---|---|---|
| United Arab Emirates | ARE | North Africa and Middle East | 1.00 | 1.15 | 1.42 | 1.00 | 66 | 0 | 1.60 | Near-frontier | 0.71 | 1.00 |
| United Kingdom | GBR | High-income | 1.58 | 2.32 | 2.15 | 2.22 | 101 | 56 | 1.61 | Regressing | 1.03 | 1.40 |
| United Republic of Tanzania | TZA | Sub-Saharan Africa | 1.57 | 1.14 | 1.00 | 1.10 | 12,157 | 1,098 | 7.62 | Stalled | 1.09 | 0.70 |
| United States of America | USA | High-income | 1.61 | 2.34 | 2.30 | 3.30 | 771 | 537 | 2.40 | Regressing | 1.44 | 2.05 |
| Uzbekistan | UZB | Central Europe, Eastern Europe, and Central Asia | 8.08 | 16.97 | 29.35 | 28.64 | 11,065 | 10,679 | 33.70 | Regressing | 0.98 | 3.55 |
| Vanuatu | VUT | Southeast Asia, East Asia, and Oceania | 2.14 | 3.42 | 2.42 | 2.86 | 32 | 21 | 5.73 | Regressing | 1.18 | 1.33 |
| Venezuela (Bolivarian Republic of) | VEN | Latin America and Caribbean | 4.08 | 4.99 | 6.03 | 7.15 | 1,199 | 1,031 | 15.23 | Regressing | 1.19 | 1.75 |
| Viet Nam | VNM | Southeast Asia, East Asia, and Oceania | 2.13 | 1.86 | 1.64 | 1.60 | 2,323 | 869 | 3.20 | Stalled | 0.98 | 0.75 |
| Yemen | YEM | North Africa and Middle East | 4.73 | 4.40 | 2.96 | 3.24 | 4,107 | 2,838 | 5.18 | Sustained catch-up | 1.09 | 0.68 |
| Zambia | ZMB | Sub-Saharan Africa | 1.56 | 2.09 | 2.30 | 2.09 | 4,764 | 2,484 | 14.47 | Regressing | 0.91 | 1.34 |
| Zimbabwe | ZWE | Sub-Saharan Africa | 1.38 | 1.22 | 1.51 | 1.45 | 3,619 | 1,127 | 10.06 | Stalled | 0.96 | 1.05 |

Note. The 137 eligible countries (2023 deaths ≥25) sum to 333,635 avoidable deaths; the text-quantified 333,803 covers all 204 countries and territories, the 168-death difference coming from low-count countries below the eligibility floor.

EFR, episode-fatality ratio (LRI deaths per incident episode); gap = country EFR / within-super-region 10th-percentile frontier EFR. Avoidable deaths = max(0, deaths − episodes × frontier EFR) at 2023. COVID ratio = gap 2023/gap 2019; long-run ratio = gap 2023/gap 1990. Eligibility: ≥25 LRI deaths in 2023; the 67 low-count countries are excluded from classification and rankings but retained in the released country-year panel. Phenotype definitions: sustained catch-up, long-run ratio ≤0.7 with no net widening by 2019; near-frontier, gap ≤1.5 at all four anchors; regressing, long-run ratio ≥1.3; all other eligible countries, stalled.

**Table S2. Priority countries, 2023: EFR gap × avoidable LRI deaths (top 15, ranked by avoidable deaths × ln gap)**

| Country | Super-region | Gap 2023 | Avoidable deaths | Observed LRI deaths |
|---|---|---|---|---|
| Nigeria | Sub-Saharan Africa | 2.40 | 67,490 | 116,482 |
| Uzbekistan | Central Europe, Eastern Europe, and Central Asia | 28.60 | 10,679 | 11,065 |
| India | South Asia | 1.60 | 54,109 | 143,192 |
| Niger | Sub-Saharan Africa | 3.90 | 18,738 | 25,191 |
| Afghanistan | North Africa and Middle East | 6.30 | 8,634 | 10,268 |
| Myanmar | Southeast Asia, East Asia, and Oceania | 6.40 | 7,006 | 8,296 |
| Indonesia | Southeast Asia, East Asia, and Oceania | 3.20 | 9,757 | 14,197 |
| Egypt | North Africa and Middle East | 4.10 | 7,649 | 10,096 |
| Democratic Republic of the Congo | Sub-Saharan Africa | 1.90 | 14,470 | 30,177 |
| Chad | Sub-Saharan Africa | 2.60 | 8,621 | 13,920 |
| Philippines | Southeast Asia, East Asia, and Oceania | 3.40 | 6,223 | 8,767 |
| Haiti | Latin America and Caribbean | 9.60 | 3,379 | 3,774 |
| Côte d'Ivoire | Sub-Saharan Africa | 2.80 | 7,246 | 11,371 |
| Mexico | Latin America and Caribbean | 8.20 | 3,344 | 3,810 |
| Azerbaijan | Central Europe, Eastern Europe, and Central Asia | 24.70 | 1,884 | 1,964 |

Avoidable deaths = max(0, deaths − episodes × frontier EFR), deterministic estimates from GBD 2023 central values; the 2023 national panel sums to the global LRI avoidable total of 333,803. Displayed avoidable deaths sum to 229,229 owing to rounding; the unrounded total is 229,230 (68.7% of the 2023 global total).

**Table S3. COVID-window movers, 2019–2023: the ten fastest catch-up countries (eligible countries)**

| Country | Super-region | Gap 2019 | Gap 2023 | Ratio |
|---|---|---|---|---|
| Botswana | Sub-Saharan Africa | 2.17 | 1.49 | 0.69 |
| United Arab Emirates | North Africa and Middle East | 1.42 | 1.00 | 0.70 |
| Turkmenistan | Central Europe, Eastern Europe, and Central Asia | 23.13 | 17.17 | 0.74 |
| Albania | Central Europe, Eastern Europe, and Central Asia | 3.72 | 2.86 | 0.77 |
| Honduras | Latin America and Caribbean | 3.37 | 2.62 | 0.78 |
| Bolivia (Plurinational State of) | Latin America and Caribbean | 6.66 | 5.32 | 0.80 |
| Gabon | Sub-Saharan Africa | 1.53 | 1.24 | 0.81 |
| El Salvador | Latin America and Caribbean | 4.28 | 3.56 | 0.83 |
| Japan | High-income | 1.38 | 1.15 | 0.83 |
| Slovakia | Central Europe, Eastern Europe, and Central Asia | 3.51 | 2.93 | 0.83 |

Ratio = gap 2023/gap 2019. Low-count countries (<25 LRI deaths in 2023) are excluded. Over the same window the global median gap rose from 1.80 (2019) to a peak of 2.00 (2021, +10.8%, computed from unrounded medians) and returned to 1.86 (2023); the 2021 peak rests on interpolated episode denominators.

**Table S4. COVID-window movers, 2019–2023: the ten largest regressing countries (eligible countries)**

| Country | Super-region | Gap 2019 | Gap 2023 | Ratio |
|---|---|---|---|---|
| Ecuador | Latin America and Caribbean | 2.87 | 4.57 | 1.59 |
| Kazakhstan | Central Europe, Eastern Europe, and Central Asia | 5.33 | 8.44 | 1.58 |
| Cambodia | Southeast Asia, East Asia, and Oceania | 3.42 | 5.03 | 1.47 |
| United States of America | High-income | 2.30 | 3.30 | 1.44 |
| Georgia | Central Europe, Eastern Europe, and Central Asia | 3.11 | 4.36 | 1.40 |
| France | High-income | 1.10 | 1.54 | 1.40 |
| Canada | High-income | 2.35 | 3.27 | 1.39 |
| Afghanistan | North Africa and Middle East | 4.63 | 6.29 | 1.36 |

| Country | Super-region | Gap 2019 | Gap 2023 | Ratio |
|---|---|---|---|---|
| Saudi Arabia | North Africa and Middle East | 1.89 | 2.54 | 1.34 |
| Peru | Latin America and Caribbean | 2.48 | 3.31 | 1.34 |

Ratio = gap 2023/gap 2019. Low-count countries (<25 LRI deaths in 2023) are excluded. Over the same window the global median gap rose from 1.80 (2019) to a peak of 2.00 (2021, +10.8%, computed from unrounded medians) and returned to 1.86 (2023); the 2021 peak rests on interpolated episode denominators.

#### Table S5. Median EFR gap to the within-super-region LRI survival frontier, by super-region and year, 1990– 2023

| Year | Central Europe, Eastern Europe, and | High-income | Latin America and Caribbean | North Africa and Middle East | South Asia | Southeast Asia, East Asia, and Oceania | Sub-Saharan Africa |
|---|---|---|---|---|---|---|---|
| 1990 | 2.13 | 1.52 | 2.67 | 2.36 | 1.09 | 1.75 | 1.57 |
| 1991 | 2.18 | 1.49 | 2.71 | 2.47 | 1.10 | 1.75 | 1.62 |
| 1992 | 2.10 | 1.45 | 2.62 | 2.54 | 1.11 | 1.76 | 1.66 |
| 1993 | 2.00 | 1.47 | 2.48 | 2.79 | 1.08 | 1.79 | 1.69 |
| 1994 | 2.19 | 1.51 | 2.30 | 2.67 | 1.13 | 1.96 | 1.62 |
| 1995 | 2.27 | 1.60 | 2.19 | 2.18 | 1.13 | 2.00 | 1.60 |
| 1996 | 2.15 | 1.59 | 2.19 | 2.27 | 1.15 | 1.99 | 1.53 |
| 1997 | 2.09 | 1.58 | 2.19 | 2.35 | 1.11 | 1.95 | 1.49 |
| 1998 | 2.18 | 1.51 | 2.14 | 2.43 | 1.11 | 1.96 | 1.50 |
| 1999 | 2.18 | 1.56 | 2.09 | 2.76 | 1.20 | 1.94 | 1.51 |
| 2000 | 2.24 | 1.51 | 2.19 | 2.84 | 1.08 | 2.07 | 1.48 |
| 2001 | 2.39 | 1.56 | 2.33 | 2.80 | 1.07 | 2.12 | 1.45 |
| 2002 | 2.36 | 1.47 | 2.38 | 2.81 | 1.06 | 2.14 | 1.46 |
| 2003 | 2.29 | 1.45 | 2.48 | 2.60 | 1.08 | 2.08 | 1.42 |
| 2004 | 2.23 | 1.60 | 2.63 | 2.50 | 1.10 | 2.01 | 1.50 |
| 2005 | 2.10 | 1.58 | 2.73 | 2.38 | 1.10 | 1.96 | 1.56 |
| 2006 | 2.06 | 1.70 | 2.89 | 2.40 | 1.10 | 1.99 | 1.54 |
| 2007 | 2.16 | 1.67 | 2.83 | 2.24 | 1.06 | 2.00 | 1.49 |
| 2008 | 2.21 | 1.74 | 2.91 | 2.08 | 1.05 | 1.98 | 1.54 |
| 2009 | 2.22 | 1.75 | 2.79 | 2.02 | 1.11 | 1.93 | 1.60 |
| 2010 | 2.30 | 1.81 | 2.90 | 1.98 | 1.12 | 1.85 | 1.67 |
| 2011 | 2.31 | 1.80 | 2.85 | 1.93 | 1.12 | 1.88 | 1.63 |
| 2012 | 2.31 | 1.86 | 2.94 | 1.90 | 1.16 | 1.84 | 1.64 |
| 2013 | 2.13 | 1.83 | 2.95 | 2.03 | 1.16 | 1.80 | 1.59 |
| 2014 | 2.19 | 1.80 | 2.91 | 2.15 | 1.14 | 1.75 | 1.62 |
| 2015 | 2.29 | 1.70 | 2.88 | 1.92 | 1.09 | 1.72 | 1.68 |
| 2016 | 2.31 | 1.65 | 2.58 | 1.59 | 1.09 | 1.70 | 1.73 |
| 2017 | 2.39 | 1.65 | 2.64 | 1.62 | 1.10 | 1.65 | 1.75 |
| 2018 | 2.28 | 1.62 | 2.57 | 1.75 | 1.13 | 1.61 | 1.75 |
| 2019 | 2.33 | 1.65 | 2.43 | 1.70 | 1.19 | 1.72 | 1.83 |
| 2020 | 2.23 | 2.01 | 2.32 | 1.61 | 1.20 | 1.93 | 1.85 |

| Year | Central Europe, Eastern Europe, and | High-income | Latin America and Caribbean | North Africa and Middle East | South Asia | Southeast Asia, East Asia, and Oceania | Sub-Saharan Africa |
|---|---|---|---|---|---|---|---|
| 2021 | 2.32 | 1.91 | 2.31 | 1.85 | 1.21 | 2.49 | 1.80 |
| 2022 | 2.68 | 1.91 | 2.21 | 1.81 | 1.24 | 2.27 | 1.77 |
| 2023 | 2.60 | 1.82 | 2.43 | 1.68 | 1.25 | 2.12 | 1.78 |

Gap = country EFR/frontier EFR, where the frontier is the 10th percentile of country EFRs within each super-region and year; the super-region median of country gaps is shown. Values for 2020–2022 rest in part on linearly interpolated episode denominators (see Methods).

### Table S6. Convergence statistics for the cross-country distribution of the EFR gap, 1990–2023

| Year | CV of gap (all countries) | SD of log gap (all | CV of gap (eligible |
|---|---|---|---|
| 1990 | 0.8871 | 0.6678 | 0.8584 |
| 1991 | 0.9193 | 0.6806 | 0.8891 |
| 1992 | 0.9300 | 0.6842 | 0.9007 |
| 1993 | 0.9292 | 0.6870 | 0.8977 |
| 1994 | 0.9716 | 0.6934 | 0.9462 |
| 1995 | 0.9867 | 0.6897 | 0.9697 |
| 1996 | 0.9848 | 0.6959 | 0.9682 |
| 1997 | 0.9937 | 0.6989 | 0.9824 |
| 1998 | 1.0299 | 0.7091 | 1.0188 |
| 1999 | 1.0085 | 0.7074 | 1.0012 |
| 2000 | 1.0125 | 0.7161 | 1.0116 |
| 2001 | 1.0292 | 0.7253 | 1.0356 |
| 2002 | 1.0245 | 0.7288 | 1.0275 |
| 2003 | 1.0268 | 0.7253 | 1.0325 |
| 2004 | 1.0246 | 0.7206 | 1.0366 |
| 2005 | 0.9994 | 0.7152 | 1.0114 |
| 2006 | 0.9878 | 0.7184 | 0.9983 |
| 2007 | 0.9921 | 0.7188 | 1.0060 |
| 2008 | 0.9903 | 0.7164 | 1.0049 |
| 2009 | 0.9697 | 0.7054 | 0.9796 |
| 2010 | 0.9783 | 0.7067 | 0.9828 |
| 2011 | 1.0063 | 0.7073 | 1.0164 |
| 2012 | 1.0208 | 0.7125 | 1.0341 |
| 2013 | 1.0303 | 0.7174 | 1.0403 |
| 2014 | 1.0520 | 0.7186 | 1.0572 |
| 2015 | 1.0517 | 0.7130 | 1.0537 |
| 2016 | 1.0578 | 0.7053 | 1.0565 |
| 2017 | 1.1088 | 0.7042 | 1.1056 |
| 2018 | 1.1367 | 0.6981 | 1.1311 |
| 2019 | 1.2207 | 0.7024 | 1.2157 |
| 2020 | 1.1256 | 0.7045 | 1.1361 |

| Year | CV of gap (all countries) | SD of log gap (all | CV of gap (eligible |
|---|---|---|---|
| 2021 | 1.1036 | 0.7108 | 1.1234 |
| 2022 | 1.1789 | 0.7169 | 1.1903 |
| 2023 | 1.1811 | 0.7222 | 1.1813 |

σ-convergence statistics: CV, cross-country coefficient of variation of the gap; SD of log gap, standard deviation of the natural logarithm of the gap. Dispersion widened between 1990 and 2023 under both measures. β-convergence (“onvergence in growth rates”): regressing the 1990–2023 change in log gap on the 1990 log gap across the 137 eligible countries gives slope -0.105 (SE 0.047; r = -0.188, p = 0.028); the corresponding levels regression (gap 2023 − gap 1990 on gap 1990) gives slope 0.113 (SE 0.083), p = 0.176. The weak β signal is consistent in part with regression to the mean and is far too weak to arrest the widening of the cross-country distribution.

**Table S7. External sensitivity checks: national mean elevation and the UHC service coverage index versus the 2023 EFR gap, 204 countries**

| Country | Gap 2023 | Mean elevation (m) | UHC service coverage index 2023 | Eligible for classification |
|---|---|---|---|---|
| Afghanistan | 6.29 | 1,792 | 42 | Yes |
| Albania | 2.86 | 663 | 71 | Yes |
| Algeria | 1.67 | 559 | 70 | Yes |
| American Samoa | 1.55 | – | – | No |
| Andorra | 2.17 | 1,258 | 75 | No |
| Angola | 2.35 | 1,062 | 44 | Yes |
| Antigua and Barbuda | 1.84 | – | 81 | No |
| Argentina | 4.57 | 697 | 80 | Yes |
| Armenia | 8.35 | 1,947 | 69 | Yes |
| Australia | 1.79 | 276 | 89 | No |
| Austria | 0.61 | 935 | 84 | No |
| Azerbaijan | 24.72 | 758 | 67 | Yes |
| Bahamas | 1.86 | 6 | 80 | No |
| Bahrain | 0.57 | 5 | 78 | No |
| Bangladesh | 1.08 | 41 | 54 | Yes |
| Barbados | 0.98 | – | 82 | No |
| Belarus | 0.71 | 160 | 80 | Yes |
| Belgium | 1.57 | 176 | 86 | No |
| Belize | 2.84 | 242 | 70 | No |
| Benin | 2.01 | 263 | 38 | Yes |
| Bermuda | 0.29 | – | – | No |
| Bhutan | 1.35 | 2,621 | 69 | Yes |
| Bolivia (Plurinational State of) | 5.32 | 1,291 | 67 | Yes |
| Bosnia and Herzegovina | 1.30 | 688 | 64 | No |
| Botswana | 1.49 | 1,038 | 60 | Yes |
| Brazil | 1.08 | 326 | 84 | Yes |
| Brunei Darussalam | 3.71 | 21 | 84 | No |
| Bulgaria | 3.47 | 409 | 73 | Yes |
| Burkina Faso | 2.27 | 302 | 47 | Yes |
| Burundi | 1.90 | 1,572 | 48 | Yes |
| Cabo Verde | 0.55 | 206 | 71 | No |
| Cambodia | 5.02 | 139 | 62 | Yes |

| Country | Gap 2023 | Mean elevation (m) | UHC service coverage index 2023 | Eligible for classification |
|---|---|---|---|---|
| Cameroon | 2.44 | 638 | 48 | Yes |
| Canada | 3.27 | 454 | 92 | Yes |
| Central African Republic | 2.91 | 594 | 39 | Yes |
| Chad | 2.63 | 519 | 26 | Yes |
| Chile | 3.28 | 1,211 | 84 | Yes |
| China | 0.50 | 1,796 | 85 | Yes |
| Colombia | 2.99 | 566 | 82 | Yes |
| Comoros | 1.58 | – | 52 | Yes |
| Congo | 1.57 | 423 | 45 | Yes |
| Cook Islands | 1.60 | – | – | No |
| Costa Rica | 1.84 | 618 | 84 | Yes |
| Croatia | 1.52 | 337 | 76 | No |
| Cuba | 1.46 | 135 | 86 | Yes |
| Cyprus | 1.27 | 355 | 76 | No |
| Czechia | 1.48 | 427 | 83 | Yes |
| Côte d'Ivoire | 2.76 | 261 | 46 | Yes |
| Democratic People's Republic of Korea | 1.75 | 590 | 77 | Yes |
| Democratic Republic of the Congo | 1.92 | 685 | 41 | Yes |
| Denmark | 1.50 | 33 | 85 | No |
| Djibouti | 1.30 | 410 | 47 | Yes |
| Dominica | 1.08 | – | 75 | No |
| Dominican Republic | 2.43 | 386 | 73 | Yes |
| Ecuador | 4.57 | 1,090 | 78 | Yes |
| Egypt | 4.13 | 308 | 71 | Yes |
| El Salvador | 3.56 | 499 | 79 | Yes |
| Equatorial Guinea | 1.40 | 525 | 49 | Yes |
| Eritrea | 1.71 | 771 | 40 | Yes |
| Estonia | 1.11 | 51 | 79 | No |
| Eswatini | 2.94 | 778 | 72 | Yes |
| Ethiopia | 1.28 | 1,250 | 33 | Yes |
| Fiji | 2.84 | 237 | 69 | Yes |
| Finland | 1.18 | 164 | 86 | No |
| France | 1.54 | 325 | 82 | Yes |
| Gabon | 1.24 | 365 | 48 | Yes |
| Gambia | 1.97 | 22 | 53 | Yes |
| Georgia | 4.36 | 1,343 | 71 | Yes |
| Germany | 0.90 | 260 | 87 | Yes |
| Ghana | 1.14 | 193 | 56 | Yes |
| Greece | 2.20 | 462 | 77 | No |
| Greenland | 14.56 | 1,701 | – | No |
| Grenada | 3.26 | – | 78 | No |
| Guam | 2.34 | – | – | No |
| Guatemala | 10.16 | 806 | 58 | Yes |
| Guinea | 2.58 | 440 | 43 | Yes |
| Guinea-Bissau | 1.96 | 34 | 43 | Yes |
| Guyana | 5.15 | 203 | 73 | Yes |

| Country | Gap 2023 | Mean elevation (m) | UHC service coverage index 2023 | Eligible for classification |
|---|---|---|---|---|
| Haiti | 9.55 | 378 | 44 | Yes |
| Honduras | 2.62 | 638 | 64 | Yes |
| Hungary | 0.81 | 162 | 80 | Yes |
| Iceland | 2.06 | 487 | 90 | No |
| India | 1.61 | 568 | 69 | Yes |
| Indonesia | 3.20 | 333 | 67 | Yes |
| Iran (Islamic Republic of) | 1.68 | 1,259 | 81 | Yes |
| Iraq | 2.17 | 303 | 64 | Yes |
| Ireland | 2.30 | 127 | 82 | No |
| Israel | 1.47 | 311 | 85 | No |
| Italy | 1.85 | 531 | 82 | Yes |
| Jamaica | 1.08 | 465 | 74 | Yes |
| Japan | 1.15 | 383 | 86 | Yes |
| Jordan | 1.71 | 750 | 74 | Yes |
| Kazakhstan | 8.44 | 351 | 83 | Yes |
| Kenya | 0.84 | 797 | 57 | Yes |
| Kiribati | 4.08 | – | 51 | No |
| Kuwait | 1.17 | 131 | 84 | Yes |
| Kyrgyzstan | 7.82 | 2,636 | 74 | Yes |
| Lao People's Democratic Republic | 8.78 | 685 | 64 | Yes |
| Latvia | 1.52 | 91 | 77 | No |
| Lebanon | 3.25 | 650 | 67 | Yes |
| Lesotho | 1.47 | 2,268 | 55 | Yes |
| Liberia | 2.35 | 225 | 49 | Yes |
| Libya | 1.67 | 415 | 71 | Yes |
| Lithuania | 1.03 | 109 | 78 | No |
| Luxembourg | 0.96 | 341 | 83 | No |
| Madagascar | 2.23 | 536 | 33 | Yes |
| Malawi | 0.93 | 890 | 52 | Yes |
| Malaysia | 1.02 | 281 | 80 | Yes |
| Maldives | 1.53 | – | 71 | No |
| Mali | 2.04 | 320 | 41 | Yes |
| Malta | 2.73 | – | 82 | No |
| Marshall Islands | 3.01 | – | 66 | No |
| Mauritania | 1.35 | 261 | 40 | Yes |
| Mauritius | 0.99 | 78 | 75 | No |
| Mexico | 8.17 | 1,040 | 79 | Yes |
| Micronesia (Federated States of) | 1.63 | – | 65 | No |
| Monaco | 2.71 | – | 85 | No |
| Mongolia | 8.48 | 1,483 | 70 | Yes |
| Montenegro | 1.42 | 1,048 | 70 | No |
| Morocco | 2.35 | 686 | 65 | Yes |
| Mozambique | 2.12 | 354 | 50 | Yes |
| Myanmar | 6.43 | 611 | 52 | Yes |
| Namibia | 1.53 | 1,085 | 66 | Yes |
| Nauru | 4.40 | – | 62 | No |

| Country | Gap 2023 | Mean elevation (m) | UHC service coverage index 2023 | Eligible for classification |
|---|---|---|---|---|
| Nepal | 1.25 | 2,101 | 58 | Yes |
| Netherlands | 1.97 | 8 | 85 | No |
| New Zealand | 2.08 | 510 | 89 | No |
| Nicaragua | 4.11 | 208 | 70 | Yes |
| Niger | 3.90 | 459 | 39 | Yes |
| Nigeria | 2.38 | 329 | 47 | Yes |
| Niue | 2.11 | – | – | No |
| North Macedonia | 1.19 | 666 | 69 | No |
| Northern Mariana Islands | 1.37 | – | – | No |
| Norway | 0.67 | 488 | 89 | No |
| Oman | 1.09 | 252 | 73 | Yes |
| Pakistan | 0.94 | 1,001 | 56 | Yes |
| Palau | 2.69 | – | 75 | No |
| Palestine | 1.26 | 211 | 65 | Yes |
| Panama | 5.47 | 262 | 82 | Yes |
| Papua New Guinea | 7.10 | 539 | 32 | Yes |
| Paraguay | 1.86 | 164 | 79 | Yes |
| Peru | 3.31 | 1,475 | 68 | Yes |
| Philippines | 3.45 | 308 | 69 | Yes |
| Poland | 0.86 | 166 | 82 | Yes |
| Portugal | 2.70 | 315 | 83 | No |
| Puerto Rico | 0.70 | 23 | – | No |
| Qatar | 0.77 | 27 | 84 | No |
| Republic of Korea | 1.04 | 286 | 88 | Yes |
| Republic of Moldova | 4.26 | 154 | 71 | Yes |
| Romania | 4.44 | 393 | 77 | Yes |
| Russian Federation | 2.60 | 349 | 81 | Yes |
| Rwanda | 1.65 | 2,021 | 59 | Yes |
| Saint Kitts and Nevis | 1.54 | – | 80 | No |
| Saint Lucia | 1.15 | 130 | 75 | No |
| Saint Vincent and the Grenadines | 2.27 | – | 80 | No |
| Samoa | 2.94 | 243 | 62 | No |
| San Marino | 1.66 | – | 74 | No |
| Sao Tome and Principe | 1.06 | – | 60 | Yes |
| Saudi Arabia | 2.54 | 642 | 83 | Yes |
| Senegal | 1.01 | 56 | 48 | Yes |
| Serbia | 1.35 | 435 | 73 | Yes |
| Seychelles | 1.60 | – | 80 | No |
| Sierra Leone | 2.97 | 173 | 48 | Yes |
| Singapore | 1.79 | – | 88 | No |
| Slovakia | 2.93 | 433 | 78 | Yes |
| Slovenia | 1.45 | 484 | 84 | No |
| Solomon Islands | 1.43 | 191 | 47 | Yes |
| Somalia | 1.86 | 280 | 30 | Yes |
| South Africa | 0.99 | 1,022 | 74 | Yes |
| South Sudan | 3.04 | 515 | 41 | Yes |
| Spain | 1.46 | 676 | 84 | Yes |

| Country | Gap 2023 | Mean elevation (m) | UHC service coverage index 2023 | Eligible for classification |
|---|---|---|---|---|
| Sri Lanka | 0.70 | 166 | 72 | Yes |
| Sudan | 3.29 | 533 | 48 | Yes |
| Suriname | 3.65 | 171 | 74 | No |
| Sweden | 1.20 | 340 | 85 | No |
| Switzerland | 1.15 | 1,360 | 87 | No |
| Syrian Arab Republic | 1.27 | 524 | 70 | Yes |
| Taiwan | 0.38 | 986 | – | Yes |
| Tajikistan | 11.20 | 3,152 | 72 | Yes |
| Thailand | 1.11 | 291 | 82 | Yes |
| Timor-Leste | 5.39 | 247 | 48 | Yes |
| Togo | 1.61 | 216 | 42 | Yes |
| Tokelau | 2.12 | – | – | No |
| Tonga | 1.53 | – | 71 | No |
| Trinidad and Tobago | 1.44 | – | 75 | No |
| Tunisia | 2.01 | 237 | 76 | Yes |
| Turkmenistan | 17.17 | 208 | 81 | Yes |
| Tuvalu | 4.17 | – | 65 | No |
| Türkiye | 1.12 | 1,156 | 77 | Yes |
| Uganda | 0.94 | 1,043 | 54 | Yes |
| Ukraine | 2.03 | 183 | 80 | Yes |
| United Arab Emirates | 1.00 | 110 | 84 | Yes |
| United Kingdom | 2.22 | 166 | 88 | Yes |
| United Republic of Tanzania | 1.10 | 998 | 49 | Yes |
| United States Virgin Islands | 0.53 | – | – | No |
| United States of America | 3.30 | 701 | 88 | Yes |
| Uruguay | 2.15 | 106 | 85 | No |
| Uzbekistan | 28.64 | 304 | 79 | Yes |
| Vanuatu | 2.86 | 293 | 52 | Yes |
| Venezuela (Bolivarian Republic of) | 7.15 | 337 | 75 | Yes |
| Viet Nam | 1.60 | 413 | 71 | Yes |
| Yemen | 3.24 | 937 | 43 | Yes |
| Zambia | 2.09 | 1,125 | 62 | Yes |
| Zimbabwe | 1.45 | 972 | 59 | Yes |

Rank correlations with the 2023 EFR gap (Spearman)

| Variable and subset | n | Spearman ρ | p value |
|---|---|---|---|
| Mean elevation, eligible countries | 135 | 0.14 | 0.107 |
| Mean elevation, all countries with data | 174 | 0.19 | 0.013 |
| UHC service coverage index, eligible countries | 136 | −0.07 | 0.417 |

| Variable and subset | n | Spearman ρ | p value |
|---|---|---|---|
| UHC service coverage index, all countries with data | 193 | −0.18 | 0.010 |

Gap = country EFR/within-super-region 10th-percentile frontier EFR at 2023. Mean elevation computed by the authors from the NOAA ETOPO1 1 arc-minute global relief model, sampled at 0.5-degree resolution and averaged over Natural Earth 1:50m country polygons; 174 of 204 panel countries matched (unmatched are small island states plus Comoros and Sao Tome and Principe). UHC service coverage index: WHO/World Bank indicator (World Bank WDI SH_UHC_SCI), 2023 values from Tracking Universal Health Coverage: 2025 Global Monitoring Report, matched for 193 of 204 countries. Eligible countries: ≥25 LRI deaths in 2023 ($n = 137$; elevation analysis $n = 135$ after excluding Comoros and Sao Tome and Principe). A Pearson check on the log gap among eligible countries gives $r = 0.124$, $p = 0.152$ ($n = 135$). Interpretation: elevation confounding does not drive the 2023 gap distribution; residual bias, if any, would inflate the gaps of high-altitude countries (Bolivia 1,291 m, Mexico 1,040 m, Ecuador 1,090 m, Afghanistan 1,792 m, Ethiopia 1,250 m). The UHC index correlates with the gap in the expected negative direction across all countries but attenuates to near zero among eligible countries, as expected because within-region frontiers themselves track system quality.
